# A Stability-Enhanced Lasso Approach for Covariate Selection in Non-Linear Mixed Effects Model

**DOI:** 10.1101/2025.05.15.25327667

**Authors:** Auriane Gabaut, Rodolphe Thiébaut, Cécile Proust-Lima, Mélanie Prague

## Abstract

Non-linear mixed effects models (NLMEMs) defined by ordinary differential equations (ODEs) are central to modeling complex biological systems over time, particularly in pharmacometrics, viral dynamics, and immunology. However, selecting relevant covariates associated with dynamics in high-dimensional settings remains a major challenge. This study introduces a novel model-building approach called Lasso-SAMBA (SAMBA: Stochastic Approximation for Model Building Algorithm) that integrates Lasso regression with a stability selection algorithm for robust covariate selection within ODE-based NLMEMs.

The method iteratively constructs models by coupling penalized regression with mechanistic model estimation using the SAEM algorithm. It extends a prior strategy named SAMBA, originally based on stepwise inclusion, by replacing this step with a penalized, stability-driven approach that reduces false discoveries and improves selection robustness. By maintaining the monotonic decrease of the information criterion through a calibrated exploration of penalization parameters, the proposed method outperforms conventional stepwise and Bayesian variable selection alternatives.

Extensive simulation studies, spanning pharmacokinetic and immunological models, demonstrate the superiority of Lasso-SAMBA in variable selection fidelity, FDR (False Discovery Proportion) control, and computational efficiency. The Lasso-SAMBA method is implemented in an R package. Applied to a Varicella-Zoster virus vaccination study, the method reveals robust, biologically plausible associations between parameters of the mechanistic model of the humoral immune response and early transcriptomic expressions. These results underscore the practical utility of our method for high-dimensional model building in systems vaccinology and beyond.

## 2. Introduction

ODE-based models, referred to as mechanistic models, are powerful tools for understanding complex biological processes. They are used in many fields such as viral dynamics [1], epidemic modeling [2], and pharmacometrics [3]. They were introduced to analyze observations repeatedly collected from multiple individuals in a population of interest [4]. These models are defined by systems of differential equations whose parameters often carry clear biological meaning—such as the half-life of a drug, the infection rate of a virus, or the turnover of immune cells. As such, they enable a causal interpretation of the relationships between compartments and offer a predictive framework that can simulate system dynamics under varying conditions [5, 6]. However, to ensure accurate predictions, it is essential to carefully identify all exogenous relationships with covariates that may influence the parameters and behavior of the model. The availability of high-dimensional data has increased significantly due to advances in data acquisition and analysis methods [7], complicating the problem of mechanistic model building. High-dimensional variable selection, where the number of covariates exceeds the number of observations, is well documented in standard regression models. However, methodological developments remain limited for its application in mechanistic models.

In a broad sense, the mechanistic model of interest belongs to the family of non-linear mixed effects models (NLMEMs), where the non-linearity does not necessarily allow for a closed-form solution and is instead defined by an ordinary differential equation (ODE). The variability inherent in the data, captured by the model parameters, can be attributed to different sources (intraindividual, interindividual, and residual). Mixed effects models on parameters of the ODE allow the study of *N* individuals exhibiting the same overall behavior. Interindividual variability in the biological parameters can be partly explained by observed covariates through fixed effects, while residual unexplained variability is captured by individual-specific random effects. This decomposition enables the model to incorporate both structured variation, due to measurable factors (e.g., demographic covariates, biomarkers), and residual variation, reflecting subject-specific deviations. Model inference and selection are essential steps in identifying the most appropriate model to best represent the data and make accurate predictions. In this article, inference methods are not seen as a major problem since many methods exist [8, 9, 10]. We mainly focus on the Stochastic Approximation version of the EM algorithm [11, 12, 13], with the selection of the “best” model among the available ones done using an extension of the Bayesian information criterion (BICc) that is well suited for NLMEMs [14]. Our main goal in this work is to propose a model-building strategy to determine which observed individual covariates have a fixed effect on the mechanistic model parameters when the set of available explanatory covariates of size *p* is high dimensional (*N* ≪ *p*).

Several approaches have been proposed in recent years to meet this challenge. The most common strategy involves a two-step process. First, the parameters of the NLME model are estimated. Then, a regression model is fitted to relate these parameters to the covariates using either a stepwise method [15] or Lasso [16], including its variations to deal with problems such as multicollinearity [17]. Bertrand et al. [18] introduced an iterative approach in which the parameter estimates are updated after being related to covariates. However, their method relied on Empirical Bayes Estimates (EBEs), which are prone to shrinkage and sensitive to study design [19]. Yuan et al. [20] proposed a correction method for shrinkage but it may fail in high-dimensional settings. An alternative to address these limitations is to integrate penalized regression into NLME models, which allows simultaneous covariate selection and parameter estimation in a one-step procedure. In this direction, Bertrand et al. [21] proposed a penalized version of the SAEM algorithm, incorporating Lasso penalization within each iteration of SAEM. However, their method does not include a penalty parameter calibration and does not support structured penalties, limiting its ability to induce structural sparsity or control model complexity using information criteria.

Recent works have extended this idea by developing more general algorithmic frameworks. Fort et al. [22] introduced a general algorithmic approach to handle intractable likelihoods through a Monte Carlo-based approximation of the gradient. This method benefits from theoretical convergence guarantees in the linear case and offers a flexible framework for incorporating various penalties. Ollier [23] proposed a proximal gradient algorithm for the simultaneous selection of covariates and correlation parameters, but its scope remains limited to moderately dimensional problems. Despite these advances, fully integrated one-step methods remain computationally intensive and difficult to tune, especially in high-dimensional settings. Since the available codes for these methods are not generic, but rather tailored to specific settings, twostep approaches remain a more practical alternative. As a more recent extension, Naveau et al. [24] proposed a Bayesian Lasso approach that controls information criteria and combines spike-and-slab priors with the SAEM algorithm. However, this work focuses on non-linear mixed effects models with closed-form outcomes. In this article, we compare our original method with the performance of their method on the published example. However, extending their approach to ODE-based NLME models remains computationally prohibitive, necessitating the development of alternative methods.

In ODE-based NLMEMs, methods for model selection such as SCM (Stepwise Covariate Modeling) [25] and COSSAC (Conditional Sampling for Stepwise Approach based on Correlation tests) [26] were proposed. SCM is a stepwise approach that systematically adds or removes covariates to optimize model fit, while COSSAC improves this process by considering correlation tests between covariates and parameter values. The SAMBA method (Stochastic Approximation for Model Building Algorithm) has been shown to be faster with similar performance [27]. In the first step, SAMBA performs an efficient screening of covariates by establishing the linear link between covariates and parameters through stochastic approximation. In the second step, it refines the selection by focusing on covariates that have been shown to be significant, thus ensuring a robust final model after several iterations of the two-step process. However, none of these methods behaves well in high-dimensional settings: the SCM or COSSAC methods are too time-consuming to be used, and the SAMBA method is based on a stepwise algorithm [28] that performs poorly in high-dimensional environments. Indeed, there is a growing risk of overfitting, multicollinearity, and the introduction of bias with the usual variable selection methods such as stepAIC. These issues emphasize the need for variable selection methods that balance model complexity and predictive power, such as penalized methods like Lasso [29, 30] within existing model-building frameworks. The method presented in this article develops a selection method for covariates in high-dimensional settings for non-linear mixed effects models by replacing the stepwise algorithm for covariate selection in complex ODE-based SAMBA with Lasso-based selection. To improve the robustness of the method, we introduce a stability selection algorithm [31, 32]. The stepwise algorithm originally used in SAMBA enforces a monotonic decrease in the information criterion across iterations, a property that Lasso does not guarantee on its own. Therefore, to control the information criterion in the new approach, we explore multiple covariate models stochastically to identify the one that minimizes the information criterion.

The paper is organized as follows. Section 3 introduces the statistical model based on a general ODE-based NLMEM and gives an overview of the SAMBA method, hereafter referred to as the step-SAMBA method, focusing on the aspects relevant to our approach. The new Lasso-SAMBA method is then presented. Simulation results are discussed in Section 4, and an application to a Varicella-Zoster virus (VZV) vaccinology study [33], revealing biological associations between parameters of the humoral immune response mechanistic model and high-dimensional transcriptomic expressions, is proposed in Section 5. Finally, in Section 6, the potential of the method is highlighted and an outlook for future work is given.

## 3. Methods

### 3.1. Non-linear mixed effects model

We consider an outcome variable, eventually multivariate,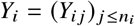 observed at time points *t*_*ij*_, with *j* indexing the occasion ( *j* = 1, …, *n*_*i*_) and *i* indexing the individual (*i* = 1, …, *N*). We assume that these observations come from an individual dynamic process *y*_*i*_ = (*f·*, ***ψ***_*i*_ ) that depends on time and on individual-specific model parameters ***ψ***_*i*_ = (*ψ*_*il*_ )_*l* ≤*m*_. These dynamics may arise from a mechanistic model defined by an ODE with parameters ***ψ***. Thus, our observations can be expressed as follows:

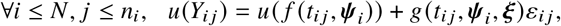

where *ε*_*ij*_ are standardized Gaussian random variables with zero mean, and *g* is a function that possibly depends on the parameters ***ξ*** and defines the residual error model. This definition is valid up to a certain predetermined link function *u*, which can be applied to account for non-linearity or other systematic deviations in the residual structure.

In the context of ODE-based NLME models, it is assumed that the individual parameters ***ψ***_*i*_ are normally distributed, or up to a transformation, around a typical population parameter ***ψ*** _*pop*_ ∈ ℝ ^*m*^, with covariance matrix **Ω**. If individual *i* is additionally characterized by covariates ***X***_*i*_ = (*X*_*i*1_, …, *X*_*ip*_ ), we can include covariate effects in the definition of the individual parameters, which leads to the following equation:

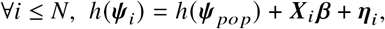

where ***β*** = ( ***β***_1_, …, ***β***_*m*_), and ***β***_*l*_ = ( *β*_*l*1_, …, *β*_*lp*_)^*T*^ represents the effect of the *p* covariates on parameter 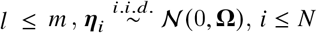, and *h* is a potential transformation function applied to the covariates or parameters.

Such models, denoted hereafter by *ℳ*, are typically estimated using the Stochastic Approximation Expectation-Maximization (SAEM) algorithm [12, 13], a stochastic variant of the Expectation-Maximization (EM) algorithm [11]. The EM algorithm aims to maximize the observed-data log-likelihood by alternating between two steps:

- an Expectation step (E-step), which computes the conditional expectation of the complete-data log-likelihood given the current estimate of the parameters,
- a Maximization step (M-step), which updates the parameters by maximizing the expected complete-data log-likelihood obtained from the E-step.

However, for complex models such as ODE-based NLME models, this conditional expectation cannot be computed analytically due to latent random effects ***η***_*i*_ . The SAEM algorithm addresses this limitation by introducing a stochastic approximation during the E-step, where individual parameters ***η***_*i*_ are sampled from their conditional posterior distribution given the current estimate of population parameters, typically using Markov Chain Monte Carlo (MCMC) methods, thus enabling parameter estimation despite the absence of a closed-form likelihood.

### 3.2. The original step-SAMBA algorithm

To select covariates in an ODE-based NLME model, the step-SAMBA [27] method creates models iteratively, starting with an empty initial model *ℳ*_0_ that contains no covariate effects or correlations, and typically assumes a combined error model *g*(*t, ψ*, ***ξ*** = (*a, b*)) = *a* + *b f* (*t, ψ*). At each iteration *k*, the algorithm updates the model *ℳ*_*k*+1_ by sequentially:

- estimating the parameters ***θ*** ^(*k*)^ = ( ***ψ*** _*po p*_, **Ω, *β, ξ***) of *ℳ*_*k*_ by maximum likelihood using the SAEM algorithm [13];
- sampling individual parameters 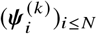 using MCMC methods;
- building three submodels with ***θ*** ^(*k*)^ and 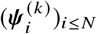: a correlation model, an error model, and a covariate model (denoted *ℳ* ^COV^).

These three submodels then form *ℳ*_*k* + 1_. A complete description of the method is available in Prague et al. [27].

Our main focus here is on the construction of the covariate part,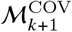. Using a single set of generated individual parameters per subject 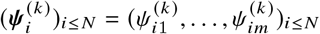, a regression model is built for each parameter ***ψ***_*l*_, where *l* ≤ *m*:

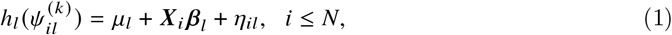

where, for *l* ≤*m, η*_*il*_ are Gaussian errors corresponding to the random effects of individual *i* on parameter *l, μ*_*l*_ is the intercept of the regression model, and ***X***_*i*_ represents the individual covariates. These are selected either by a stepwise selection procedure if there are more than 10 covariates, or by an exhaustive search among all possible models if there are fewer than 10 covariates.

The SAMBA method is an efficient approach to improve the model-building process in ODE-based NLMEMs, as demonstrated in simulation and real-world examples [27]. We propose to replace the stepwise algorithm for selecting covariates in step-SAMBA with a refined Lasso-based selection method in order to improve the variable selection process in high-dimensional data (*N* ≪ *p*).

An alternative penalization strategy would be to replace the Lasso with an Elastic Net regression [34]. Compared to the Lasso, the Elastic Net is particularly beneficial in situations with highly correlated covariates, as it combines an *L*_1_ and an *L*_2_ penalty to encourage both sparsity and the grouping of correlated variables. This extension has also been investigated, and the details are provided in Supplementary Material Section A.1.

### 3.3. The stability-enhanced Lasso-SAMBA algorithm

Similar to the step-SAMBA method, our approach performs variable selection independently for each parameter *ψ*_*l*_ in the ODE-based model. At each iteration *k* ∈ℕ, the first two steps remain unchanged: the Lasso-SAMBA method first estimates the population parameters and samples the individual parameters. Then, in the Lasso-SAMBA method, we estimate coefficients in Equation (1) with a Lasso regression [29] given a penalty parameter *λ*_*l*_:

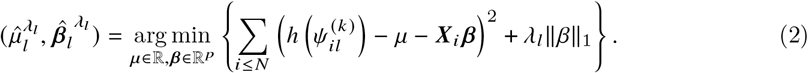

For simplicity, we now omit the explicit subscript for each parameter *l* in the remainder of the paper; for example, in Equation (2), 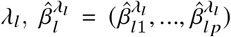 are later denoted by *λ*, 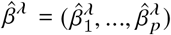.

The main goal is to identify, for each parameter of the mechanistic model, the true set *𝒮* = { *c* ≤*p*; *β*_*c*_ ≠ 0} corresponding to the selection of truly relevant covariates. For a fixed *λ*, this set can be derived, for example, by:

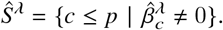

Usually in Lasso regression, the penalty parameter, and thus the selected set of covariates, is determined through cross-validation. However, this approach can result in unstable selections, particularly in high-dimensional settings [35, 31]. As an alternative, the penalty parameter can be calibrated using an information criterion, such as the BIC, by selecting the value that minimizes the criterion over a grid of candidate penalty parameters. This approach was also investigated alongside the Lasso-SAMBA algorithm, with details provided in Supplementary Material Section A.2.

To address this, we rely on a Stability Selection Algorithm [31, 32], a method developed to improve the performance of variable selection algorithms. It has been proven that stability selection achieves consistency in variable selection [31]. The central idea of the stability selection algorithm is to repeat the variable selection process a large number of times for a fixed value of *λ* on data samples that are half the size of the original dataset. A covariate *c* ≤ *p* is selected when its selection frequency across batches, denoted as 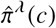, exceeds a given threshold *π* ∈ [0, 1].

In our case, we draw uniformly *n*_*SS*_ random subsamples of individuals 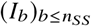 from {1, …, *N* }, each of size ⌊*N*/2⌋. For each batch *b* ≤ *n*_*SS*_, we denote 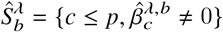 as the set of selected covariates resulting from the Lasso regression presented in Equation (2) in batch *b* with penalty parameter *λ >* 0. The final selection set for the hyperparameters (*λ, π*) is then:

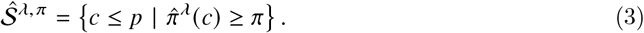

where 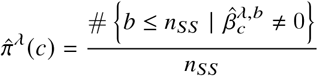 the selection frequency of covariates *c* across batches. Ultimately, this method is based on two critical hyperparameters for each parameter of the mechanistic model: *λ >* 0 and *π* ∈ [0, 1].

### 3.4. Calibration of parameters (*λ, π*) and BICc

In the step-SAMBA method, the selection of covariates by a stepwise algorithm seems to ensure a reduction of the criterion used for model construction. Consequently, the models *ℳ*_*k*_ constructed during the iterations of the algorithm have a BICc criterion that usually decreases as the iterations progress, allowing the algorithm to stop at a certain point. However, when using a Lasso method, the decrease of the BICc may be lost. This behavior appears to result from the decrease in the BIC (Bayesian Information Criterion) of the individual parameter model (1) during the stepwise covariate selection at each iteration of the step-SAMBA method. To preserve this property, we conduct an exploration step for selecting the covariate model *ℳ*^*COV*^ during the covariate model building step of each Lasso-SAMBA iteration *k*. We explore a set Λ × Π of hyperparameters (*λ, π*) that results in models *ℳ*^*COV*^ (*λ, π*), among which the one minimizing the BIC is selected as the candidate covariate model *ℳ*^*COV*^ . The covariate model is then chosen as 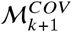 if it has a lower BIC than the previous model 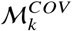; otherwise, 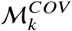 is retained. The schematic pipeline of the Lasso-SAMBA procedure is presented in Figure 1.

**Figure 1:**
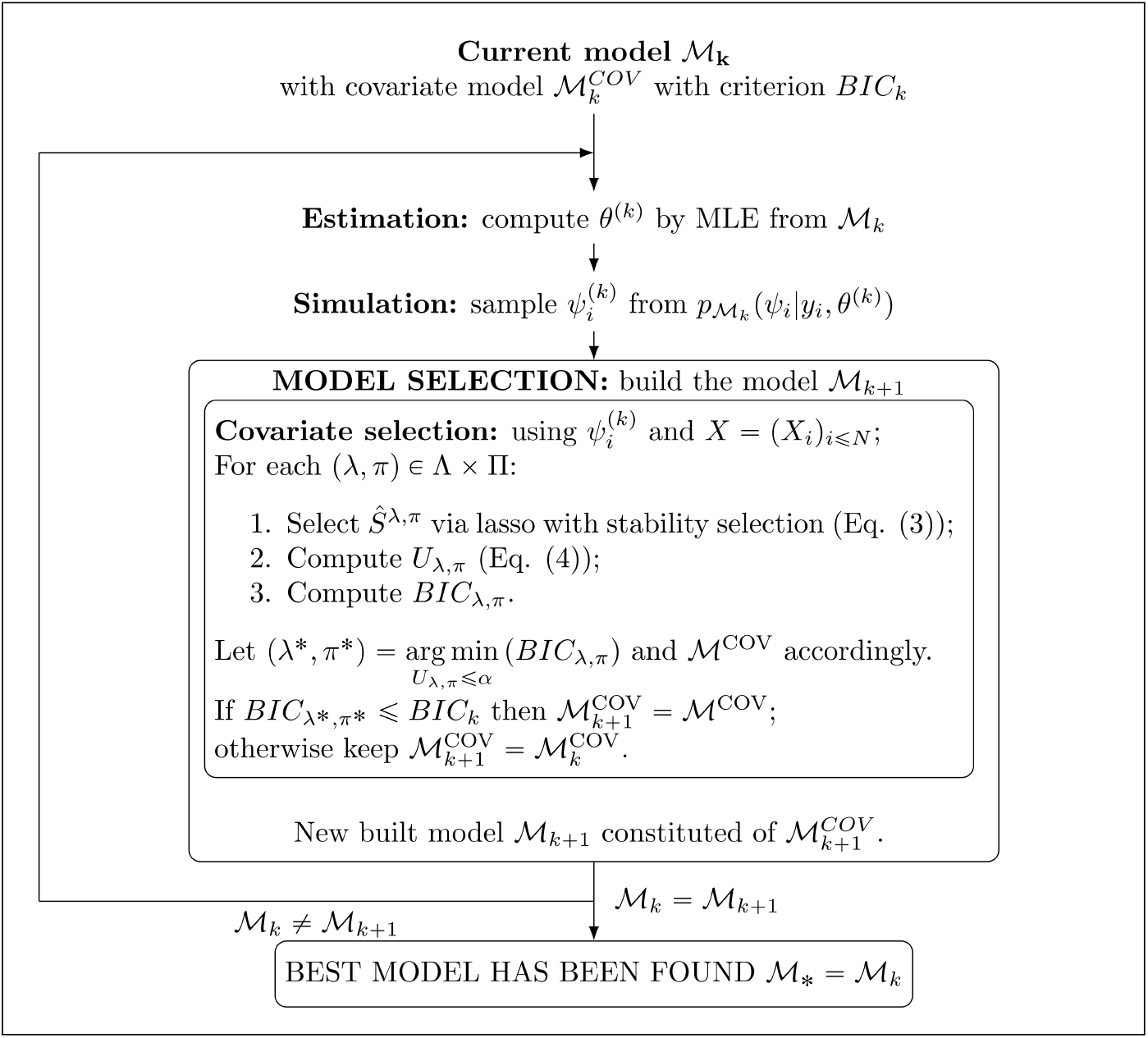
Lasso-SAMBA method pipeline.

In other words, rather than identifying a single set of parameters (*λ, π*), we aim to explore a space that yields several covariate models, searching for one that minimizes the BIC.

To characterize a good set Λ × Π, we use the upper bound for the expected number of false discoveries derived by Meinshausen and Bühlmann [31]. If we denote by *V* ^*λ, π*^ the set of falsely selected variables in 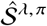, we have, for *π* ∈]0.5; 1]:

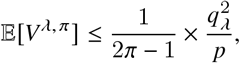

where 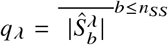 is the average number of features that are selected at least once by the Lasso selection with penalty *λ*. This upper bound for the expected number of false discoveries can then be transformed into an upper bound for the expected proportion of false discoveries, the False Discovery Rate (FDR):

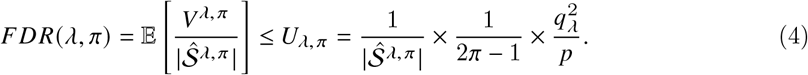

Starting from a space of *λ* ∈Λ, which ranges from dense to sparse models, as is usually done [36], and Π =]0.5; 1], we compute 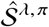 for each *λ* ∈ Λ, *π* ∈ Π and the induced upper bounds *U*_*λ, π*_ . We first remove from Λ×Π any set of parameters that does not fulfill the condition *U*_*λ, π*_ ≤ *α* for a fixed hyperparameter *α* ∈ [0; 1]. For the remaining set of parameters Λ × Π\{(*λ, π*), *U*_*λ, π*_ *> α*}, we choose the final set of selected covariates such that:

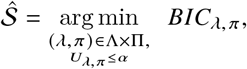

where *BIC*_*λ, π*_ is the information criterion of the linear model in Equation (1) with 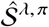 as the selected covariates.

In practice, the grids Λ and Π must be chosen to balance exploration of dense versus sparse models while ensuring computational feasibility. Following common recommendations [37] and implementations of variable selection methods [38], Λ is defined so that the resulting model sizes range from empty models to approximately min (*p, N* / 2) selected covariates, where *N* is the number of individuals. For the threshold parameter, we consider a grid Π ranging from 0.5 to 1, as suggested by theoretical results on stability selection [31], with fine increments to explore.

Alternatively, instead of generating batches from a single set of individual parameters per subject, we can sample multiple datasets of individual parameters from the posterior distribution. The methodology detailed below still applies, with each batch of data simply replaced by a set of simulated individual parameters. This approach has also been investigated, and details are provided in Supplementary Material Section A.3.

### 3.5. Implementation

The new method is implemented in the LSAMBA R [39] package, available at https://github.com/aurianegbt/LSAMBA, using the Rsmlx [27, 40] and lixoftConnectors [41] packages as a base. For the Lasso estimation in Lasso-SAMBA, we use the sharp package [38], which implements stability-based variable selection. All code was run with Lixoft Monolix 2023R1.

## 4. Simulation studies

In this section, we evaluate the performance of the Lasso-SAMBA method in terms of covariate selection through simulation studies. We conduct a simulation study under two distinct frameworks to assess the performance of the proposed method in sparse settings. The aim is to compare the newly developed procedure with step-SAMBA [27] and SAEMVS [24] in terms of variable selection accuracy. The first has been described above in Section 3.2, while the latter consists of iteratively applying a SAEM procedure, where the E-step estimates posterior inclusion probabilities of variables using Monte Carlo sampling, and the M-step updates model parameters by maximizing a penalized likelihood, effectively shrinking irrelevant coefficients toward zero for sparse model recovery. We calibrate both reference methods using the hyperparameters and initialization strategies detailed in the original studies. For our Lasso-SAMBA, the only hyperparameter requiring specification is the constraint on the expected number of false discoveries, which we fixed at 10%. A sensitivity analysis over a range of values is reported in Supplementary Material Section D (in Supplementary Figures 23 and Supplementary Table 10). Data are generated using biologically motivated models presented in each subsection: a toy pharmacokinetics model (Section 4.2) and a humoral immune response model (Section 4.3). For each framework, 100 independent replicates are generated. In addition to these reference methods, we also investigate several alternative strategies, including the use of an Elastic Net penalty [34](see Section 3.2), the calibration of the penalty parameter via BIC (see Section 3.3), and the sampling of multiple sets of individual parameters per subject for the stability selection algorithm (see Section 3.4). The results of these complementary analyses are detailed in Supplementary Material Section A.

### 4.1. Performance evaluation

In the context of covariate selection, a true positive (TP) is a covariate truly associated with the parameter and correctly identified as such; a false positive (FP) is not associated but incorrectly selected; a false negative (FN) is truly associated but missed; and a true negative (TN) is not associated and correctly excluded. The overarching goal is to minimize false discoveries.

In particular, for our method, the aim is to reduce the False Discovery Rate (FDR) without significantly increasing the False Negative Rate (FNR), defined as:

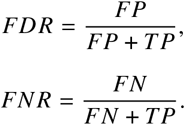

To assess the performance of the model, we compare both the FDR and FNR, while also considering the *F*_1_-score, which provides a single metric that balances the trade-off between FP and FN:

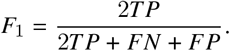

### 4.2. A toy pharmacokinetic model

We replicate the simulation study provided in Naveau et al. [24]. We simulate a one-compartment pharmacokinetic model with linear elimination. The model describes the concentration *y*_*ij*_ of a drug in the effect compartment measured at 12 time points *t* _*j*_, *j* = 1, …, 12, for individual *i* with *i* = 1, …, 200:

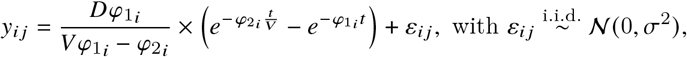

and

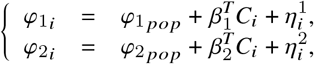

where 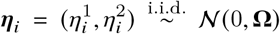 are random effects explaining individual residual heterogeneity, and *σ*^2^ is the variance of measurement error. Time points for all individuals are *t* _*j*_ ∈{0.05, 0.15, 0.25, 0.4, 0.5, 0.8, 1, 2, 7, 12, 24, 40 }. The administered dose *D* = 100 and the apparent volume of distribution *V* = 30 are known constants. The individual parameters are the individual-specific rate constant *φ*_1_ and *φ*_2_. These parameters are to be estimated and related to individual-specific covariates.

For each individual *i*, the covariate matrix *C*_*i*_ ∈ℝ^500^ is simulated from independent binomial distributions with success probability 0.2. The covariates are standardized. The effect parameters (*β*_1_, *β*_2_ ) are such that covariates 1-3 are associated with 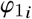, and covariates 3-5 are associated with 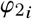, leaving 495 covariates not associated with any parameter. The parameter values are detailed in Supplementary Material Section B.

In this initial simulation scenario, designed to evaluate variable selection under controlled conditions, we allow both correlation and covariate model building through the Lasso-SAMBA and step-SAMBA methods in order to compare results with the SAEMVS algorithm. Here, we observe that the Lasso-SAMBA method markedly outperforms step-SAMBA with respect to FDR control. The median FDR achieved by step-SAMBA is 88% (95% CI: 79%, 92%), while Lasso-SAMBA reduces this to a striking 0.0% (95% CI: 0.0%, 14.3%), as summarized in Table 1. This considerable improvement in FDR does not come at the expense of reduced power.

**Table 1:** False Discovery Rate (FDR), False Negative Rate (FNR), and *F*_1_-score computed for each simulation framework, comparing the original step-SAMBA, the Lasso-SAMBA, with an error control threshold of 10%, and the SAEMVS methods. (NA: not applied; NC: non convergent).

| Metric | PK model<br>binomial covariates<br>$N = 200, p = 500$ | Vaccinology model<br>Gaussian covariates<br>$N = 100, p = 200$ | Vaccinology model<br>Gaussian covariates<br>$N = 100, p = 1000$ | Vaccinology model<br>non-Gaussian covariates<br>$N = 100, p = 200$ |
| --- | --- | --- | --- | --- |
| **FDR (%) :** |  |  |  |  |
| - step-SAMBA | 87.5 [78.8;91.7] | 72.7 [50.0;83.8] | NC | 70.0 [25.0;81.2] |
| - Lasso-SAMBA | 0.0 [0.0;14.3] | 0.0 [0.0;25.0] | 0.0 [0.0;32.9] | 0.0 [0.0;25.0] |
| - SAEMVS | 0.0 [0.0;100.0] | NA | NA | NA |
| **FNR (%) :** |  |  |  |  |
| - step-SAMBA | 0.0 [0.0;0.0] | 0.0 [0.0;0.0] | NC | 0.0 [0.0;0.0] |
| - Lasso-SAMBA | 0.0 [0.0;0.0] | 0.0 [0.0;0.0] | 0.0 [0.0;0.0] | 0.0 [0.0;0.0] |
| - SAEMVS | 0.0 [0.0;100.0] | NA | NA | NA |
| **$F_1$-score (%) :** |  |  |  |  |
| - step-SAMBA | 22.2 [15.4;34.9] | 42.9 [27.9;66.7] | NC | 46.2 [31.6;85.7] |
| - Lasso-SAMBA | 100.0 [92.3;100.0] | 100.0 [85.7;100.0] | 100.0 [80.1;100.0] | 100.0 [85.7;100.0] |
| - SAEMVS | 100.0 [0.0;100.0] | NA | NA | NA |

Indeed, as illustrated in Figure 3, panel A, which plots covariate selection frequencies across 100 replicated datasets, Lasso-SAMBA retains the same number of true positive selections as step-SAMBA, demonstrating that the improved FDR is achieved without sacrificing sensitivity. Consequently, the *F*_1_-score shows a substantial increase: from 22% (95% CI: 15%, 35%) with step-SAMBA to 100% (95% CI: 92%, 100%) with Lasso-SAMBA.

Further evaluation of model selection quality is presented in Figure 2, panel A. This figure shows the proportion of exact model recoveries and models that strictly include the correct covariates. The step-SAMBA method fails to recover the exact model in any replicate. In contrast, Lasso-SAMBA successfully recovers the exact model structure in 96% of simulations. Notably, neither method yields final models with false negative covariates, indicating consistent inclusion of all relevant associations.

**Figure 2:**
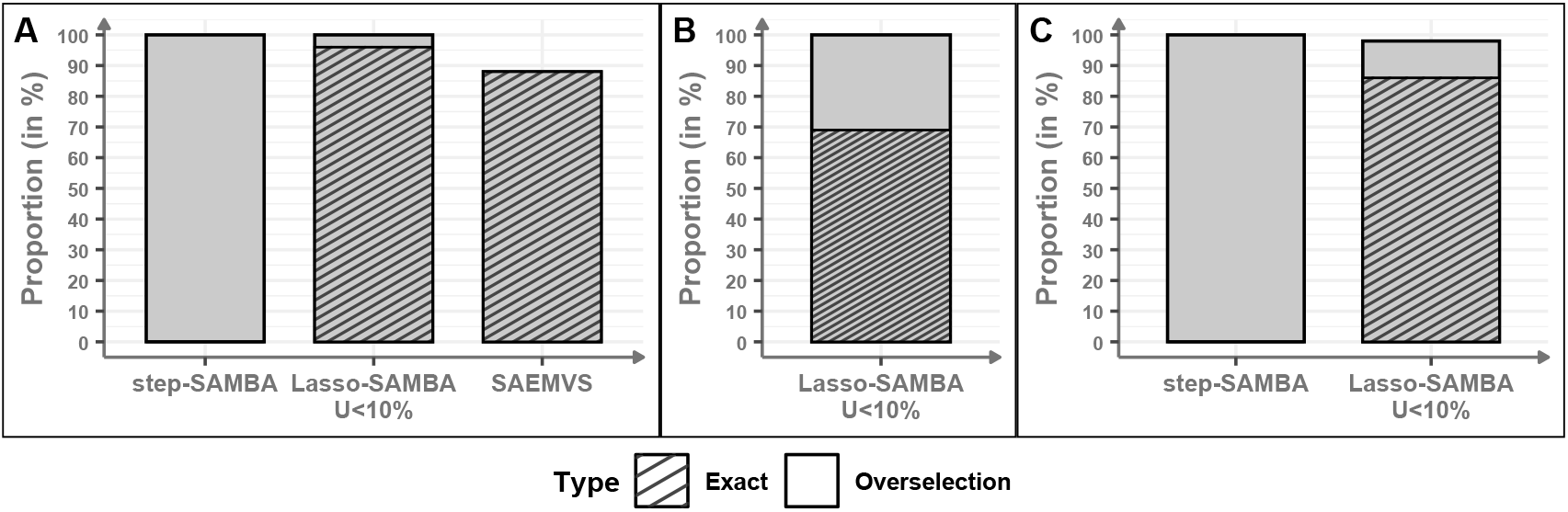
Proportion of exact models (no false positives or false negatives) and models that strictly include the exact model (no false negatives but with false positives) by step-SAMBA, the Lasso-SAMBA, with an error control threshold of 10%, and the SAEMVS methods. (A) Pharmacokinetic model with categorical covariates (*N* = 200, *p* = 500); (B) Vaccinology framework with Gaussian-correlated covariates (*N* = 100, *p* = 1000); (C) Vaccinology framework with randomly drawn correlated covariates (*N* = 100, *p* = 200).

**Figure 3:**
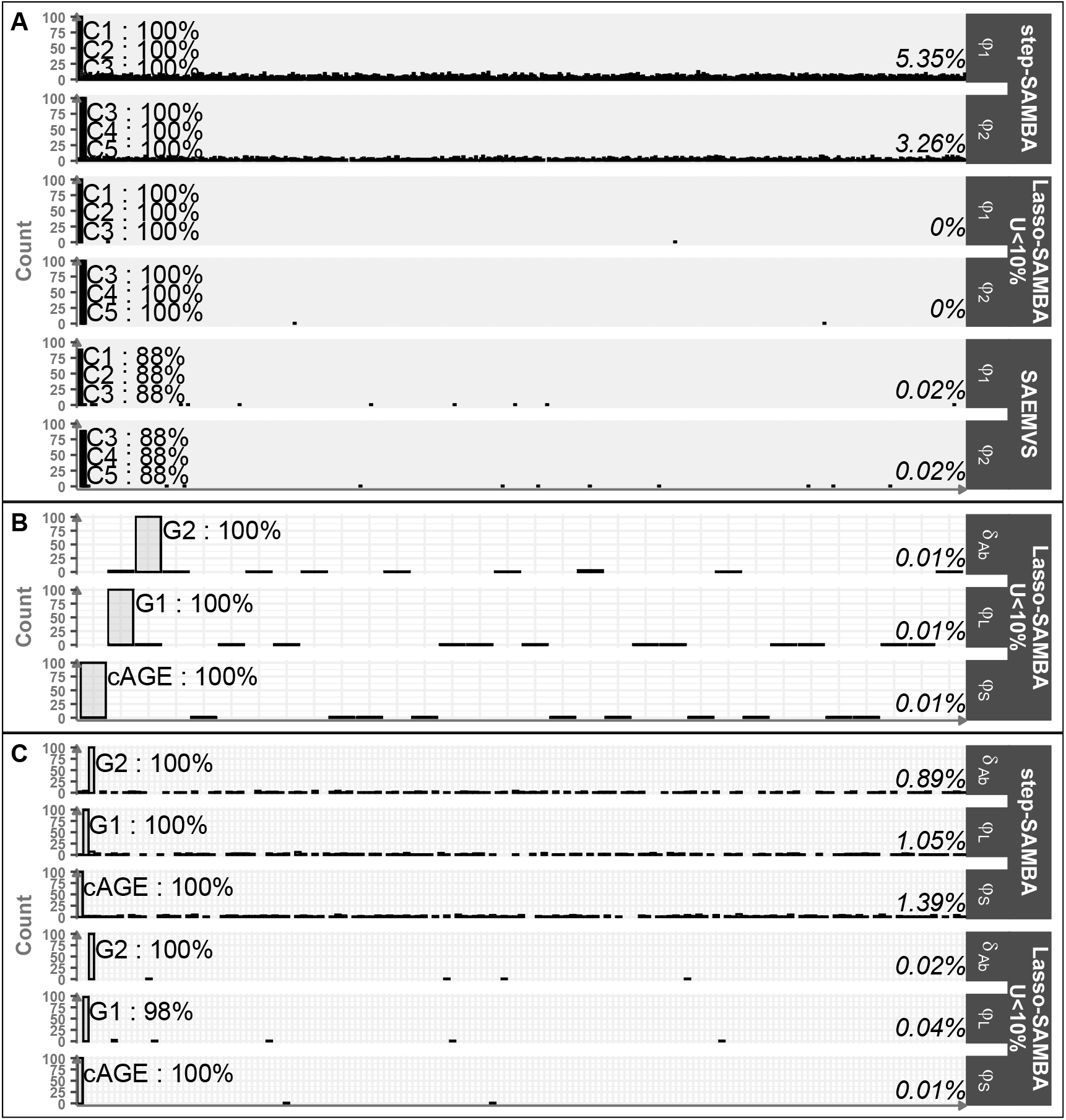
Covariate selection frequency across simulation frameworks for the step-SAMBA, the Lasso-SAMBA, with an error control threshold of 10%, and the SAEMVS methods. The mean selection frequency of false discoveries is shown on the right side of each histogram. (A) Pharmacokinetic model with categorical covariates (*N* = 200, *p* = 500); (B) Vaccinology framework with Gaussian-correlated covariates (*N* = 100, *p* = 1000); (C) Vaccinology framework with randomly drawn correlated covariates (*N* = 100, *p* = 200).

We also compare the performance of Lasso-SAMBA with the SAEMVS method. In this context, SAEMVS achieves exact model recovery in 88% of replicates. However, unlike Lasso-SAMBA, which maintains zero false negatives across all replicates, SAEMVS occasionally omits relevant variables. Table 1 also indicates substantial variability in performance, as the *F*_1_-score can be as low as 0% in some replicates.

Besides the main comparison with step-SAMBA and SAEMVS, we also evaluated alternative strategies described in Supplementary Material Section A. The version using an Elastic Net penalty provides nearly identical results to the stability-based approach, with equivalent FDR and *F*_1_-score values and comparable runtimes, confirming the robustness of the proposed procedure to the choice of penalization. In contrast, the version calibrated via BIC exhibits markedly higher false discovery rates, lower *F*_1_-scores, and longer computation times, mainly due to the additional iteration needed. These complementary results, detailed in Supplementary Sections A.1 and A.2 respectively, further support the advantages of the stability-based calibration adopted in Lasso-SAMBA.

### 4.3. Simulation of humoral immune response

To emulate a realistic biological process relevant to our real-data application [42], we simulate antibody kinetics for 100 individuals based on a mechanistic model incorporating two types of antibody-secreting cells (ASCs): short-lived (S) and long-lived (L), which secrete antibodies (Ab) at rates *φ*_*S*_ and *φ*_*L*_, respectively, and are characterized by their distinct half-lives *δ*_*S*_ and *δ*_*L*_. Antibody concentrations decay over time according to a rate parameter *δ*_*Ab*_. The dynamics are driven by an ordinary differential equation as follows:

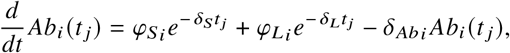

with individual parameters

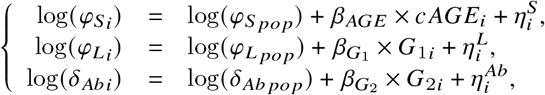

with 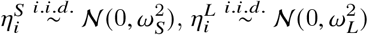 and 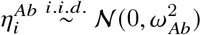, and ( *β*_*AGE*_, *β*_*G*_, *β*_*G*_ ) the effect parameters. For each individual, the centered age *c AGE* follows a normal distribution with variance 16, and *G*_1_ and *G*_2_ are drawn from standardized normal distributions. The process is observed at times *t* _*j*_ ∈{ 0, 7, 21, 123, 180, 300 }.

To simulate a high-dimensional setting, 200 covariates are generated by adding 197 noise variables to the 3 informative ones. Two data generation scenarios are considered to test the robustness of the methods: one involving purely Gaussian covariates and another with a heterogeneous mix of randomly drawn distributions, including both categorical and continuous variables. Besides these two scenarios with *p* = 200 covariates, we also investigated a higher-dimensional setting with *p* = 1000 Gaussian-correlated covariates. In all cases, correlation is introduced among the covariates to mimic realistic dependencies observed in biomedical datasets. Full details of the covariate generation process are provided in Supplementary Material Section B.

In the Gaussian-correlated simulation framework, covariate model building was performed using both Lasso-SAMBA and step-SAMBA. The Lasso-SAMBA method again demonstrates a marked improvement in FDR over the step-SAMBA method. The median FDR decreases from 73% (95% CI: 50%, 84%) to 0.0% (95% CI: 0.0%, 25.0%), as shown in Table 1. Supplementary Figure 26, panel A, confirms that this improvement in specificity is not achieved at the cost of sensitivity, as the number of true discoveries remains comparable between the two methods. In turn, the *F*_1_-score is drastically increased under Lasso-SAMBA, reaching a median of 100% (95% CI: 86%, 100%). Moreover, the 95% confidence intervals for both FDR and *F*_1_-score do not overlap between the methods. Supplementary Figure 26, panel B, further illustrates that, whereas the step-SAMBA method fails to recover the exact model in any replicate, Lasso-SAMBA identifies the correct model structure in 81% of cases. In the Gaussian-correlated simulation framework with *p* = 1000 covariates, step-SAMBA is unusable: the underlying stepwise routine repeatedly fails to fit the regression model. In contrast, Lasso-SAMBA remains stable and accurate: as summarized in Table 1, the median FDR is 0.0% (95% CI: 0.0%, 32.9%) and the median *F*_1_-score reaches 100% (95% CI: 80.1%, 100%). Again, as highlighted by Figure 3, panel B, no false negatives are observed in any replicate. As shown in Figure 2, panel B, Lasso-SAMBA recovers the true model in 69% of replicates.

To assess the robustness of the method to deviations from classical assumptions, we investigate a simulation setting where covariates are randomly drawn from heterogeneous distributions, including categorical and non-Gaussian continuous variables. Even under these more complex and less idealized conditions, Lasso-SAMBA continues to substantially outperform the benchmark method. The median FDR drops from 70% (95% CI: 25%, 81%) to 0.0% (95% CI: 0%, 25%), as summarized in Table 1. Figure 3, panel C, indicates that the number of true discoveries remains largely unaffected, while the median *F*_1_-score increases from 46% (95% CI: 31%, 85%) to 100% (95% CI: 88%, 100%). Model selection quality is again improved: as shown in Figure 2, panel C, step-SAMBA fails to identify the true model in any replicate, while Lasso-SAMBA achieves exact recovery in 86% of cases.

Finally, a comparative analysis of computational times across simulation frameworks, when comparison was possible, reveals that Lasso-SAMBA is generally faster. Although individual iterations of Lasso-SAMBA tend to be slower due to the optimization procedure, the algorithm converges in fewer steps compared to step-SAMBA. In the two more complex vaccinology-inspired frameworks with *p* = 200 covariates, Lasso-SAMBA requires a median of two iterations to converge, versus four for step-SAMBA. In the vaccinology-inspired frameworks with *p* = 1000 covariates, comparison was not available as the step-SAMBA method fails to run. The Lasso-SAMBA method, however, successfully converges in a median of 30 713 seconds (approximately 8.5 hours). These runtime differences are displayed in Supplementary Figure 25.

Along with the main analyses, we also assessed the alternative strategies described in Supplementary Material Section A. In the Gaussian and heterogeneous vaccinology frameworks, the version using an Elastic Net penalty yields results nearly identical to those of the stability-based approach, with comparable FDR, *F*_1_-scores, and runtimes, confirming the robustness of the method to the penalization scheme. Conversely, the BIC-calibrated version leads to higher false discovery rates, lower *F*_1_-scores, and slightly longer runtimes, consistent with the trends observed in the pharmacokinetic scenario. Finally, the version based on repeated sampling of individual parameters achieves reasonable performances but it is markedly slower, with runtimes exceeding three hours on average compared to less than five minutes for the stability-based approach. Detailed results for these complementary analyses are reported in Supplementary Sections A.1, A.2 and A.3, respectively.

## 5. Application to VZV vaccination data

To illustrate the utility of our approach in a real-world setting, we apply the proposed methodology to a publicly available dataset from a clinical study of immune response to vaccination against the Varicella-Zoster virus (VZV). This dataset, accessible under the accession number SDY984 on the ImmPort platform [43], stems from the study titled “Zoster vaccine in young and elderly,” and includes gene expression and antibody response data following immunization with ZOSTAVAX, a live attenuated vaccine targeting VZV [33].

ZOSTAVAX [44] is employed both for the primary prevention of varicella—a childhood illness marked by viremia, fever, and vesicular skin lesions—and for the prevention of herpes zoster, a reactivation of latent VZV that manifests as a localized dermatomal rash. In this study, 35 adult volunteers were administered the vaccine, with longitudinal sampling of whole blood at baseline and days 1, 3, 7, 14, 30, 90, and 180 post-vaccination. Transcriptomic data were obtained at days 0, 1, 3, and 7, while antibody levels were measured at days 0, 7, 14, 30, 90, and 180. Of the 10,086 profiled genes, a subset of 784 protein-coding genes was selected based on functional annotation with roles in interferon signaling, type I interferon response, neutrophil activation, inflammation, cytokine/chemokine activity, and cell cycle regulation, as defined by Biomart [45] and Li et al. [46].

To characterize antibody kinetics post-vaccination, we use the humoral immune response model proposed by Pasin et al. [42], which was studied in simulations in Section 4.3 and is detailed in Supplementary Material Section C. The analysis specifically explored which early gene expression signatures are predictive of early humoral responses, focusing on the first day post-vaccination—a time point previously shown to exhibit maximal transcriptomic variation. We investigate the relationship between these genes and the parameters *φ*_*S*_, *φ*_*L*_, and *δ*_*S*_ using the step-SAMBA and Lasso-SAMBA methods.

The gene associations identified by the step-SAMBA and Lasso-SAMBA methods are presented in Table 2. Step-SAMBA identified 15 genes, while the Lasso-SAMBA method yielded a sparse model in which LEP and KIFC1 were uniquely associated with the model parameters *φ*_*S*_ and *φ*_*L*_, respectively. The model resulting from the model-building procedure for parameters *φ*_*S*_ and *φ*_*L*_ respectively follows:

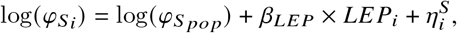

and

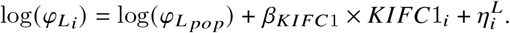

**Table 2:** Results from the step-SAMBA and the Lasso-SAMBA methods for the ZOSTAVAX vaccine. The initial model had an empty covariate structure, a constant error model, and no correlation. Both methods started from the same initial conditions, with covariate model selection being the only difference.

| Parameter | Genes selected |  |
| --- | --- | --- |
|  | step-SAMBA | Lasso-SAMBA |
| $\varphi_S$ | ASPM, CDC45, **LEP**, NOTCH1, SP110 | LEP |
| $\varphi_L$ | GDPD5, IKZF2, **KIFC1**,<br>PSORS1C1, SLC16A5, SLC50A1, SRP72 | KIFC1 |
| $\delta_S$ | ISTNA1, RAB2A, SLC31A1 | |

The final model estimates, incorporating these genes, are described in detail in Supplementary Material Section C. These findings implicate LEP and KIFC1 as candidate biomarkers of antibody-secreting cell dynamics during early vaccine-induced immunity. LEP and KIFC1 both belong to the cell cycle pathway. While the first encodes the leptin hormone, the second encodes kinesin family member C1. Their potential role in immunity is discussed in Section 6.

To evaluate model specificity and control for spurious associations, we conduct model selection using a null covariate dataset without any biological signal. This artificial dataset was generated by fitting a multivariate Gaussian distribution to the empirical gene expression values of the 784 preselected genes. Under these conditions, the Lasso-SAMBA method did not identify any significant associations with the antibody model parameters, supporting its robustness to spurious associations. In contrast, the step-SAMBA method produced six false-positive associations—one involving *φ*_*S*_ and five involving *φ*_*L*_.

Further assessment of model stability is perform via nonparametric bootstrap analysis using 500 resampled datasets (Figure 4). The Lasso-SAMBA method consistently identified LEP and KIFC1 as the most stable covariates, with selection frequencies of 25.2% and 22.6%, respectively. The step-SAMBA method also frequently selected LEP (45.8%) and KIFC1 (46.2%) but exhibited greater model variability; no other genes surpassed a selection frequency of 12.8%. Additionally, step-SAMBA failed to converge in 7 out of 500 iterations, primarily due to overfitting-related estimation issues.

**Figure 4:**
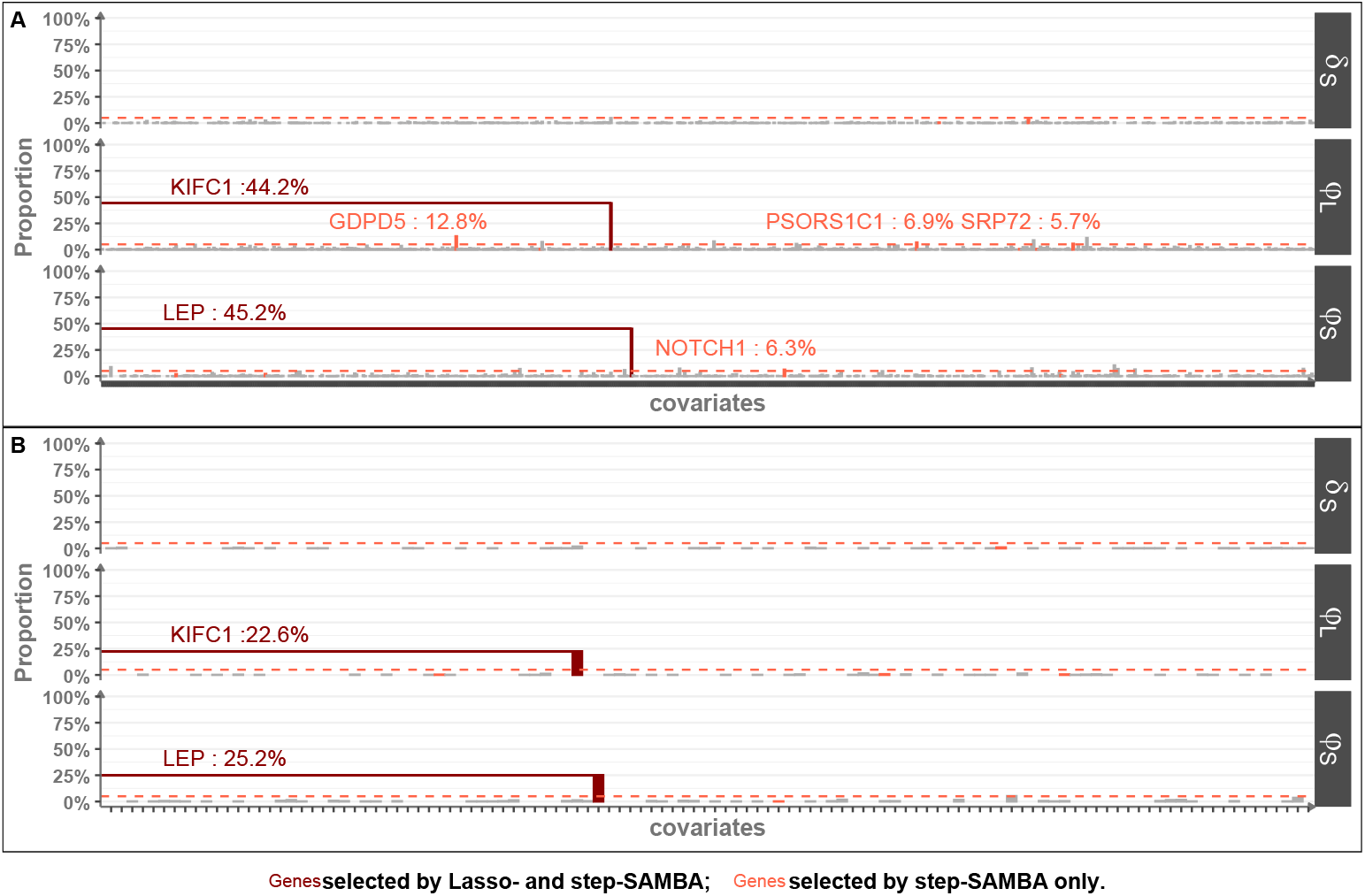
Bootstrap selection frequencies for covariates across all parameter models. The top panel shows results for the step-SAMBA method (493 successful runs), and the bottom panel for the Lasso-SAMBA method (500 runs). Selection frequencies above 5% of selected genes by any of the procedures are indicated.

Together, these findings underscore the enhanced robustness and reproducibility of the Lasso-SAMBA method in identifying biologically plausible gene-parameter associations in the context of humoral immune modeling.

## 6. Discussion

Model selection remains a central challenge in the development of reliable statistical models, particularly within the context of personalized medicine, where the accurate identification of individual-specific covariates can substantially influence therapeutic decision-making. In this paper, we proposed a method for selecting covariates in high-dimensional settings by integrating a Lasso approach for covariate selection within the framework of non-linear mixed effects models described by ordinary differential equations. Although the method was presented for a univariate longitudinal outcome variable, it also applies to any ODE-based model with multivariate longitudinal data.

The motivation for this enhancement stems from the limitations of traditional stepwise selection procedures as implemented in step-SAMBA which, although computationally tractable, are known to be susceptible to overfitting and the inclusion of non-informative covariates, especially in high-dimensional settings [47]. By integrating the Lasso, coupled with a stability selection procedure, we aimed to improve the selection of relevant covariates while preserving computational feasibility, in contrast to the approach by Naveau et al. [24].

Our simulation studies demonstrated a marked improvement in the control of FDR compared to other procedures, without compromising FNR. This improvement was consistent across diverse pharmacokinetic and vaccinology-inspired model scenarios. These findings reinforce prior conclusions on the robustness of penalized regression approaches in complex hierarchical models [48]. In terms of dimensionality, our simulation studies explored scenarios with up to p=1000 covariates and N=200 individuals. We found that the Lasso-SAMBA approach remained computationally feasible in these high-dimensional settings, with increased runtime. In fact, in our simulation, beyond 1000 covariates the step-SAMBA method became impractical, underscoring the scalability advantage of our penalized selection method. Even though larger covariate counts are conceptually addressable, they would likely require significantly more computation time due to the costly ODE-based model fitting at each iteration.

Furthermore, in the real-data application involving antibody response to Varicella-Zoster virus vaccination, the Lasso-SAMBA approach exhibited superior performance in identifying biologically plausible biomarkers while producing a sparser covariate selection. This is consistent with the detection of true signals observed in our simulation studies. The LEP gene, through leptin, promotes humoral immunity by enhancing B cell proliferation, survival, and antibody production, playing a critical role in supporting adaptive immune responses during infection and vaccination [49, 50]. In contrast, KIFC1 encodes a mitotic motor protein essential for ensuring chromosomal stability during rapid cell proliferation in cancer and immunotherapie [51, 52]. Further exploratory analysis investigating the effect of LEP and KIFC1 on humoral dynamics is required to confirm their relevance. KIFC1 also has the ability to promote stable mitotic spindle formation during early B cell development, where centriole duplication is frequent but must be tightly regulated [53]. These results suggest that our method improves the reliability of gene-covariate relationships in biomedical research involving ODE-based modeling.

Despite these promising results, certain modeling assumptions should be acknowledged. In our approach, we assumed that the random effects and residual errors follow normal distributions, a standard assumption in non-linear mixed-effects models that facilitates identifiability and tractable inference [54]. However, this assumption may not always hold in practice. Studies have shown that deviations from normality, especially in the random effects distribution, can inflate variance component estimates and, in more severe cases, bias fixed effects estimates [55]. Nevertheless, estimation remains relatively robust under moderate deviations, particularly for fixed effects and could be diagnosed by looking at the eta-shrinkage parameter estimation. Another important consideration is potential model misspecification. The reliability of covariate selection and parameter inference inherently depends on the correctness of the assumed model structure. If the underlying ODE or the covariate-parameter relationship is misspecified - a common challenge in complex mechanistic modeling - even a sophisticated selection procedure may identify spurious covariates or miss relevant ones.

The methodological integration we propose holds promise for broader applications, particularly in fields such as pharmacogenomics and systems pharmacology, where model interpretability and variable selection are of paramount importance. An appealing feature of our approach is that it relies on a single, interpretable hyperparameter—the target false discovery rate controlled at each iteration. Its sensitivity was assessed across multiple simulation settings for thresholds of 5%, 10%, and 20%, revealing no substantial deterioration in performance, with only a slight increase in false positives at higher thresholds. In future work, increasing the granularity of this calibration could help refine sensitivity assessment and provide practical guidance for choosing this parameter. Future investigations could extend this approach to non-linear relationships between covariates and parameters. Moreover, an extension using group penalties (such as group Lasso [56] or sparse group Lasso [57]) could enable covariate selection at a pathway or gene-set level. This would be especially useful in applications like transcriptomics or pathway analysis, where covariates can be grouped by biological function or signaling pathway. By selecting groups of related covariates rather than individual genes, the model could identify entire biological pathways associated with the parameters, providing higher-level interpretability in contexts such as gene expression data. Another perspective for future work is to extend the framework to time-varying parameters and time-varying covariates, allowing dynamic effects of biomarkers or treatments on model parameters and improving applicability to longitudinal personalized medicine studies. This would however require substantial changes since the proposed methodology requires individual parameters to be time-invariant. We leave this for future research.

## Data Availability

Code for running the simulations and applications are available at

https://github.com/aurianegbt/LassoSAMBA

## Abbreviations

ASC: Antibody-Secreting Cell
CI: Confidence Interval
EM: Expectation-Maximization
FDP: False Discovery Proportion
FDR: False Discovery Rate
FNR: False Negative Rate
MCMC: Markov Chain Monte Carlo
NLMEM: Non-Linear Mixed Effects Model
ODE: Ordinary Differential Equation
SAEM: Stochastic Approximation Expectation-Maximization
SAMBA: Stochastic Approximation for Model Building Algorithm
VZV: Varicella-Zoster virus

## Data and code availability

Code for running the simulations and applications is available at https://github.com/aurianegbt/LassoSAMBA.

## Acknowledgments

This study was carried out within the framework of the University of Bordeaux’s France 2030 program RRI PHDS. We thank the Human Immunology Project Consortium for providing the ImmPort platform for open-access immunology data. We also thank the investigators who contributed data for study SDY984, supported by the Human Immunology Project Consortium (HIPC) RFA-AI-15-041.

We are grateful to Simulations Plus, Lixoft division, for granting free academic access to the MonolixSuite. Computational resources for this study were provided by the PlaFRIM (Plateforme Fédérative pour la Recherche en Informatique et Mathématiques).

We disclose that generative AI tools were used to polish the English language.

## Supporting information

Additional supporting information may be found in the online version of the article at the publisher’s website.

