## Supplementary material for "A Stability-Enhanced Lasso Approach for Covariate Selection in Non-Linear Mixed Effects Model": Suplementary Material

This supplementary material provides additional details and results supporting the main manuscript. It includes (A) alternative methodological strategies, including BIC-based penalty calibration, an elastic net extension, and the use of multiple posterior samples of individual parameters; (B) the full specification of simulation models and parameters; (C) the mathematical formulation and estimation of the ODE-based model used in the real-data application; and (D) complementary analysis figures such as sensitivity assessments, computational benchmarking, and convergence diagnostics.

The method is implemented in the LSAMBA R [1] package, available at `https://github.com/aurianegbt/LSAMBA` and in CRAN. Codes for running the simulations and applications are available at `https://github.com/aurianegbt/LassoSAMBA`. All code was run with Lixoft Monolix 2023R1.

### A. Alternative methodological strategies

#### A.1. Elastic net extension

##### A.1.1. Method

In the following, covariate selection is performed within the regression framework linking the sampled individual parameters to the observed covariates, as defined in Equation (1) of the main manuscript and reproduced here:

$$h_l(\psi_{il}^{(k)}) = \mu_l + \boldsymbol{X}_i\boldsymbol{\beta}_l + \eta_{il}, \quad i \leq N, \tag{S.1}$$

where, for $l \leq m$, $\eta_{il}$ are Gaussian errors corresponding to the random effects of individual $i$ on parameter $l$, $\mu_l$ is the intercept of the regression model, and $\boldsymbol{X}_i$ represents the individual covariates. The objective is then to identify the subset of covariates with non-zero coefficients in $\boldsymbol{\beta}_l$, corresponding to those that significantly explain the variability of parameter $\psi_l$ across individuals.

In this alternative specification, the covariate selection step relies on the Elastic Net [2] rather than on the Lasso. At each iteration $k$, after estimation of the population parameters and sampling of the individual parameters $(\boldsymbol{\psi}_i^{(k)})_{i\leq N}$, each parameter $\psi_l$ of the mechanistic model is related to the covariates through the penalized regression problem:

$$(\hat{\mu}_l^{\lambda,\alpha}, \hat{\boldsymbol{\beta}}_l^{\lambda,\alpha}) = \underset{\mu\in\mathbb{R},\boldsymbol{\beta}\in\mathbb{R}^p}{\arg\min} \left\{ \sum_{i\leq N} \left( h(\psi_{il}^{(k)}) - \mu - \boldsymbol{X}_i\boldsymbol{\beta} \right)^2 + \lambda \left( \alpha\|\beta\|_1 + \tfrac{1-\alpha}{2}\|\beta\|_2^2 \right) \right\}, \tag{S.2}$$

where $\lambda > 0$ determines the overall penalization level and $\alpha \in [0,1]$ controls the compromise between the $L_1$ and $L_2$ components. The Lasso corresponds to $\alpha = 1$, while ridge regression is recovered when $\alpha = 0$.

As in the main manuscript, we now omit the explicit subscript for each parameter $l$ in the remainder of the paper; for example, in Equation (S.2), $(\lambda_l, \alpha_l, \hat{\beta}_l^{\lambda_l,\alpha_l})$ are later denoted by $(\lambda, \alpha, \hat{\beta}^{\lambda,\alpha})$.

For the Elastic Net specification, later called ELasticNet-SAMBA, the calibration step follows the same stability-selection strategy as described for Lasso-SAMBA (see Section 2.4). Jointly tuning the three hyperparameters $(\lambda, \pi, \alpha)$ would substantially increase computational costs, as it would require exploring a much larger grid. Therefore, in the present work we fixed the mixing parameter to $\alpha = 0.5$, which balances the $L_1$ and $L_2$ components, and focused the calibration on $(\lambda, \pi)$. We also evaluated the sensitivity of the results to this choice of $\alpha$.

##### A.1.2. Results for the ElasticNet-SAMBA method

We compared the ElasticNet-SAMBA to the stability-based Lasso-SAMBA across three simulation frameworks considered in the main manuscript: the pharmacokinetic (PK) model with $N = 200$ individuals and $p = 500$ covariates, the vaccinology model with Gaussian correlated covariates ($N = 100$, $p = 200$), and the vaccinology model with heterogeneous correlated covariates ($N = 100$, $p = 200$) (see Section 3.2 and 3.3).

Overall, ElasticNet-SAMBA with $\alpha = 0.5$ provided results that were remarkably close to those of Lasso-SAMBA across all simulation frameworks. In the PK model ($N = 200$, $p = 500$), the two methods produced nearly indistinguishable outcomes, with ElasticNet-SAMBA yielding a median FDR of 0.0% (95% CI: 0.0%, 7.5%) and an $F_1$-score of 100.0% (95% CI: 96.0%, 100.0%), compared to 0.0% (95% CI: 0.0%, 14.3%) and 100.0% (95% CI: 92.3%, 100.0%) for Lasso-SAMBA, as reported in Supplementary Table 1. In the Gaussian vaccinology scenario ($N = 100$, $p = 200$), ElasticNet-SAMBA performed almost as well, with a single false negative observed across all replicates, whereas Lasso-SAMBA recovered all relevant covariates. Nevertheless, FDR remained controlled at 0.0% (95% CI: 0.0%, 29.4%), and the $F_1$-score matched that of Lasso-SAMBA. In the heterogeneous vaccinology setting, both approaches again produced nearly identical results, with zero false negatives and a median FDR of 0.0% (95% CI: 0.0%, 25.0%).

Selection frequencies are shown in Supplementary Figure 3, which confirms this strong agreement: ElasticNet-SAMBA systematically retained the truly associated covariates and exhibited a very low rate of spurious selections. Exact model recovery, displayed in Supplementary Figure 1, was also comparable, with ElasticNet-SAMBA recovering the true covariate structure in 97% of replicates for the PK model, 78% in the Gaussian vaccinology framework, and 84% in the heterogeneous framework, versus 96%, 81%, and 86% for Lasso-SAMBA, respectively. These marginal differences underline the robustness of ElasticNet-SAMBA in high-dimensional settings.

Runtime comparisons are displayed in Supplementary Figure 2. Across all scenarios, ElasticNet-SAMBA was at least as efficient as Lasso-SAMBA, with sensibly identical runtimes: 7,070s versus 7,231s in the PK framework, 280s versus 281s in the Gaussian case, and 2,113s versus 2,109s in the heterogeneous case.

Finally, the sensitivity of ElasticNet-SAMBA to the choice of $\alpha$ was investigated in the Gaussian vaccinology framework, as shown in Supplementary Figure 4 and in Supplementary Table 2. Varying $\alpha$ between 0.1 and 0.9 produced consistent results, with FDR and FNR remaining equal to 0.0%. Confidence interval bounds exhibited the expected trends: the upper limit of the FDR interval decreased gradually from 40.0% to 25.0% as $\alpha$ increased, while the lower bound of the $F_1$-score interval rose from 66.7% to 85.7%. These results are coherent with the respective roles of ridge and Lasso penalizations: ridge improves stability of estimation, whereas Lasso is more effective for covariate selection. In practice, performance was highly stable for all $\alpha > 0.5$, showing that ElasticNet-SAMBA is robust to the choice of mixing parameter and only marginally sensitive in the small-$\alpha$ regime.

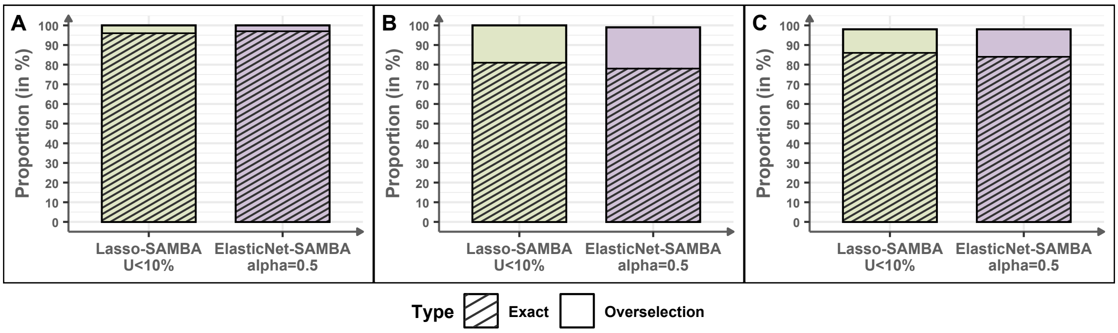

Supplementary Figure 1: Proportion of exact models (no false positives or false negatives) and models that strictly include the exact model (no false negatives but with false positives) by the Lasso-SAMBA method, with the error control threshold set at 10%, and ElasticNet-SAMBA method, with $\alpha = 0.5$. (A) Pharmacokinetic model with categorical covariates ($N = 200$, $p = 500$); (B) Vaccinology framework with Gaussian-correlated covariates ($N = 100$, $p = 200$); (C) Vaccinology framework with randomly drawn correlated covariates ($N = 100$, $p = 200$).

Supplementary Table 1: False Discovery Rate (FDR), False Negative Rate (FNR), and $F_1$-score computed for each simulation framework, comparing the step-SAMBA, the Lasso-SAMBA, with an error control threshold of 10%, and the ElasticNet-SAMBA, with $\alpha = 0.5$, methods.

| **Metric** | **PK model** binomial covariates $N = 200,\ p = 500$ | **Vaccinology model** Gaussian covariates $N = 100,\ p = 200$ | **Vaccinology model** non-Gaussian covariates $N = 100,\ p = 200$ |
|---|---|---|---|
| **FDR (%) :** | | | |
| - step-SAMBA | 87.5 [78.8;91.7] | 72.7 [50.0;83.8] | 70.0 [25.0;81.2] |
| - Lasso-SAMBA | 0.0 [0.0;14.3] | 0.0 [0.0;25.0] | 0.0 [0.0;25.0] |
| - ElasticNet-SAMBA | 0.0 [0.0;7.5] | 0.0 [0.0;29.4] | 0.0 [0.0;25.0] |
| **FNR (%) :** | | | |
| - step-SAMBA | 0.0 [0.0;0.0] | 0.0 [0.0;0.0] | 0.0 [0.0;0.0] |
| - Lasso-SAMBA | 0.0 [0.0;0.0] | 0.0 [0.0;0.0] | 0.0 [0.0;0.0] |
| - ElasticNet-SAMBA | 0.0 [0.0;0.0] | 0.0 [0.0;0.0] | 0.0 [0.0;0.0] |
| **$F_1$-score (%) :** | | | |
| - step-SAMBA | 22.2 [15.4;34.9] | 42.9 [27.9;66.7] | 46.2 [31.6;85.7] |
| - Lasso-SAMBA | 100.0 [92.3;100.0] | 100.0 [85.7;100.0] | 100.0 [85.7;100.0] |
| - ElasticNet-SAMBA | 100.0 [96.0;100.0] | 100.0 [80.1;100.0] | 100.0 [82.7;100.0] |

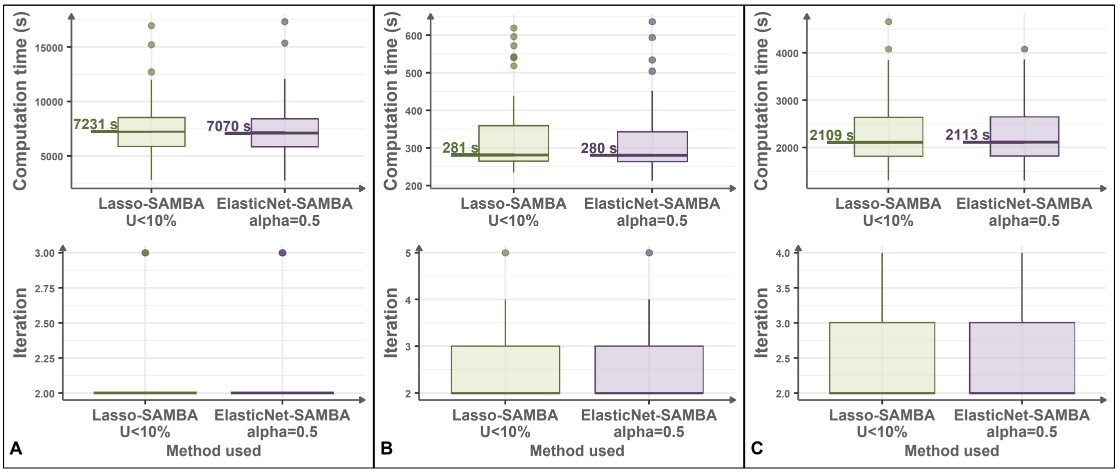

Supplementary Figure 2: Comparison of computation time distributions across simulation frameworks, for the Lasso-SAMBA method, with the error control threshold set at 10%, and ElasticNet-SAMBA method, with $\alpha = 0.5$. (A) Pharmacokinetic model with categorical covariates ($N = 200$, $p = 500$); (B) Vaccinology framework with Gaussian-correlated covariates ($N = 100$, $p = 200$); (C) Vaccinology framework with randomly drawn correlated covariates ($N = 100$, $p = 200$).

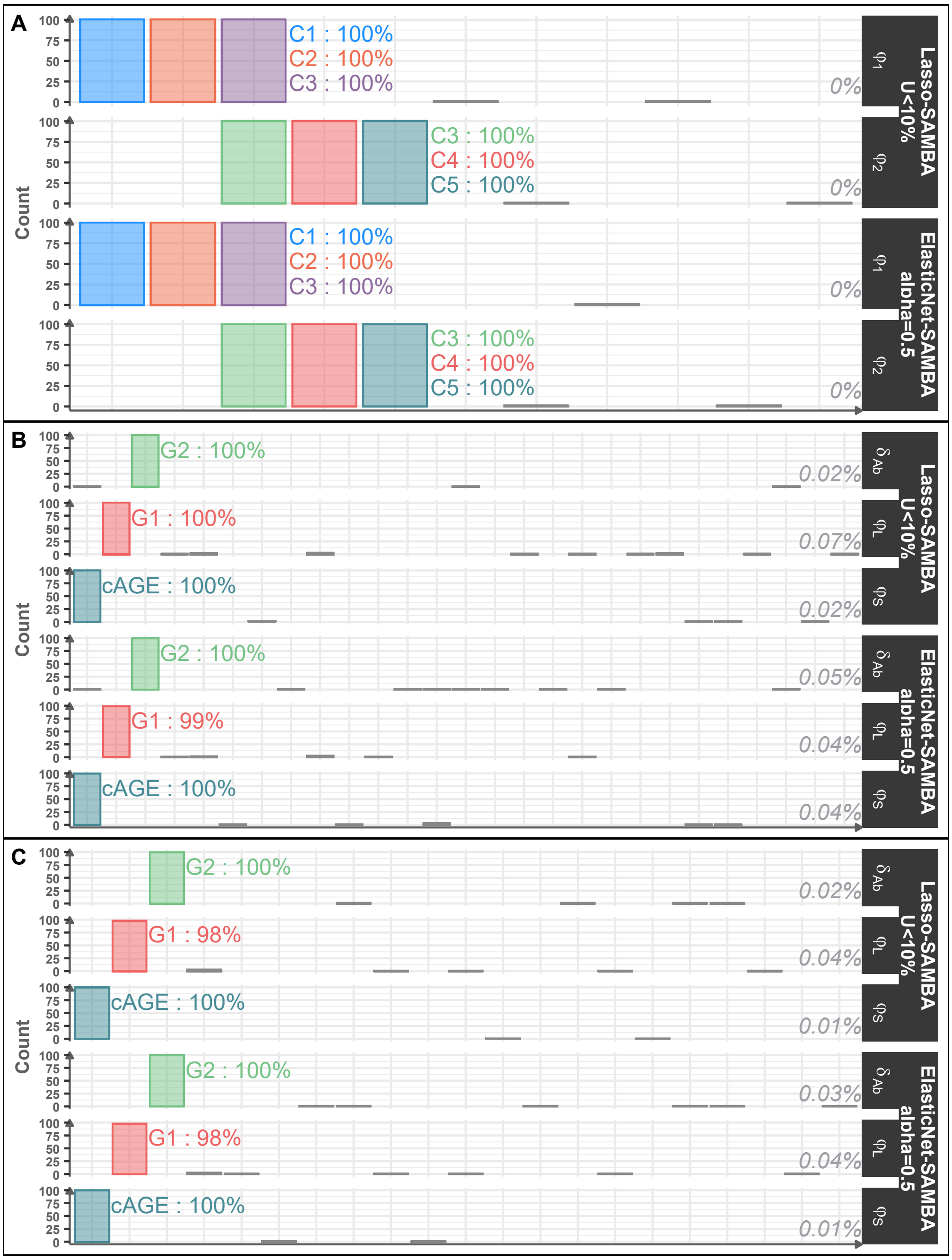

Supplementary Figure 3: Covariate selection frequency across simulation frameworks for the Lasso-SAMBA method, with the error control threshold set at 10%, and ElasticNet-SAMBA method, with $\alpha = 0.5$. The mean selection frequency of false discoveries is shown on the right side of each histogram. (A) Pharmacokinetic model with categorical covariates ($N = 200$, $p = 500$); (B) Vaccinology framework with Gaussian-correlated covariates ($N = 100$, $p = 200$); (C) Vaccinology framework with randomly drawn correlated covariates ($N = 100$, $p = 200$).

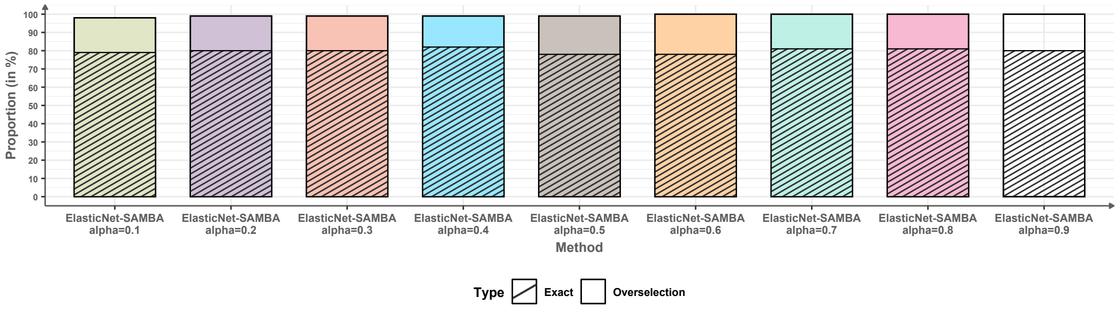

Supplementary Figure 4: Proportion of exact models (no false positives or false negatives) and models that strictly include the exact model (no false negatives but with false positives) for the ElasticNet-SAMBA method under different penalty parameter $\alpha$ in the Vaccinology framework with Gaussian-correlated covariates ($N = 100$, $p = 200$).

| $\alpha$ | **FDR (%)** | **FNR (%)** | **F1-score (%)** |
|---|---|---|---|
| 0.1 | 0.0 [0.0;40.0] | 0.0 [0.0;0.0] | 100.0 [66.7;100.0] |
| 0.2 | 0.0 [0.0;36.8] | 0.0 [0.0;0.0] | 100.0 [75.0;100.0] |
| 0.3 | 0.0 [0.0;36.8] | 0.0 [0.0;0.0] | 100.0 [75.0;100.0] |
| 0.4 | 0.0 [0.0;29.4] | 0.0 [0.0;0.0] | 100.0 [80.1;100.0] |
| 0.5 | 0.0 [0.0;29.4] | 0.0 [0.0;0.0] | 100.0 [80.1;100.0] |
| 0.6 | 0.0 [0.0;25.0] | 0.0 [0.0;0.0] | 100.0 [85.7;100.0] |
| 0.7 | 0.0 [0.0;32.9] | 0.0 [0.0;0.0] | 100.0 [80.1;100.0] |
| 0.8 | 0.0 [0.0;25.0] | 0.0 [0.0;0.0] | 100.0 [85.7;100.0] |
| 0.9 | 0.0 [0.0;25.0] | 0.0 [0.0;0.0] | 100.0 [85.7;100.0] |

Supplementary Table 2: False Discovery Rate (FDR), False Negative Rate (FNR), and $F_1$-score computed for each ElasticNet-SAMBA penalty parameter $\alpha$ in the Vaccinology framework with Gaussian-correlated covariates ($N = 100$, $p = 200$).

#### A.2. BIC-based penalty calibration

##### A.2.1. Method

Similar to the main Lasso-SAMBA framework, variable selection is performed independently for each parameter $\psi_l$ in the mechanistic model. At each iteration $k$, the population parameters are first estimated, and individual parameters $(\boldsymbol{\psi}_i^{(k)})_{i\leq N}$ are sampled. Then, for each parameter $\psi_l$, a Lasso regression is fitted as in Equation (2) from the main manuscript, tagged here (S.3) :

$$(\hat{\mu}_l^\lambda, \hat{\boldsymbol{\beta}}_l^\lambda) = \underset{\mu\in\mathbb{R},\boldsymbol{\beta}\in\mathbb{R}^p}{\arg\min} \left\{ \sum_{i\leq N} \left( h\left(\psi_{il}^{(k)}\right) - \mu - \boldsymbol{X}_i\boldsymbol{\beta} \right)^2 + \lambda\|\beta\|_1 \right\}. \tag{S.3}$$

Again, for notational simplicity, we later drop the explicit index $l$ in $\lambda_l$, $\hat{\beta}_l^{\lambda_l} = (\hat{\beta}_{l1}^{\lambda_l}, ..., \hat{\beta}_{lp}^{\lambda_l})$ and simply write $\lambda$, $\hat{\beta}^\lambda = (\hat{\beta}_1^\lambda, ..., \hat{\beta}_p^\lambda)$.

For each candidate penalty parameter $\lambda$ in a predefined grid $\Lambda$ [3], the set of selected covariates is defined as

$$\hat{S}^\lambda = \{c \leq p \mid \hat{\beta}_c^\lambda \neq 0\}.$$

We then calibrate the optimal penalty parameter by minimizing the Bayesian Information Criterion (BIC) of the regression model in Equation (S.1). The chosen covariate set is therefore $\hat{S}^{\lambda^*}$ such that:

$$\lambda^* = \arg\min_{\lambda\in\Lambda} BIC_\lambda,$$

with $BIC_\lambda$ the information criterion of the linear model in Equation (S.1) with $\hat{\mathcal{S}}^\lambda$ as the selected covariates.

This procedure is repeated independently for each parameter $\psi_l$, yielding the covariate model $\mathcal{M}_{k+1}^{COV}$ at iteration $k+1$. Similarly as the main method Lasso-SAMBA, to preserve the information-criterion monotonicity, the newly proposed model $\mathcal{M}_{k+1}^{COV}$ is only retained if its BIC is lower than that of the previous model $\mathcal{M}_k^{COV}$; otherwise, the previous model is conserved.

The BIC-based calibration preserves the step-SAMBA structure while replacing stepwise selection with a penalized regression. It is computationally efficient and requires no additional hyperparameters, since the penalty parameter $\lambda$ is directly chosen by minimizing the BIC. This approach is later called LassoBIC-SAMBA.

##### A.2.2. Simulation studies results for the LassoBIC-method

We again compared the BIC-calibrated version of our procedure, LassoBIC-SAMBA, to the stability-based Lasso-SAMBA across three simulation frameworks considered in the main manuscript: the pharmacokinetic (PK) model with $N = 200$ individuals and $p = 500$ covariates, the vaccinology model with Gaussian correlated covariates ($N = 100$, $p = 200$), and the vaccinology model with heterogeneous correlated covariates ($N = 100$, $p = 200$) (see Section 3.2 and 3.3).

Figure 7 summarizes covariate selection frequencies across 100 replicates. In all three frameworks, LassoBIC-SAMBA maintained zero false negatives, indicating that all relevant covariates were consistently retained. However, the method more frequently selected spurious covariates compared to Lasso-SAMBA, leading to substantially higher false discovery rates. As reported in Supplementary Table 3, the median FDR under LassoBIC-SAMBA reached 40.0% (95% CI: 0.0%, 64.7%) in the PK framework, 40.0% (95% CI: 0.0%, 66.7%) in the Gaussian vaccinology framework, and 25.0% (95% CI: 0.0%, 57.1%) in the heterogeneous framework, compared to 0.0%

for Lasso-SAMBA in all cases. This deterioration in specificity translated into lower $F_1$-scores: 75.0% (95% CI: 52.2%, 100.0%), 75.0% (95% CI: 50.0%, 100.0%), and 85.7% (95% CI: 60.0%, 100.0%) for LassoBIC-SAMBA versus 100% for Lasso-SAMBA. In all settings, false negative rates remained equal to 0.0% for both methods.

Model recovery quality, illustrated in Supplementary Figure 5, further emphasizes these differences. LassoBIC-SAMBA identified the exact underlying model in only 21% of replicates in the PK framework, 25% in the Gaussian vaccinology framework, and 34% in the heterogeneous framework, while Lasso-SAMBA achieved 96%, 81%, and 86%, respectively. These results confirm that, while LassoBIC-SAMBA avoids false negatives, its tendency to overselection drastically reduces the probability of exact recovery.

Finally, Figure 6 compares computation times. In the PK framework, median runtime was substantially higher for LassoBIC-SAMBA (11 820s) compared to Lasso-SAMBA (7 231s), as the BIC-calibrated procedure required more iterations to converge (median of three versus two). In the vaccinology scenarios, runtimes were closer (308s vs. 281s for Gaussian covariates, and 2 647s vs. 2 109s for heterogeneous covariates), but LassoBIC-SAMBA still tended to be slower overall. These findings indicate that, despite its conceptual simplicity and computational efficiency at the calibration step, LassoBIC-SAMBA is less efficient in practice due to increased iterations and yields systematically poorer covariate selection performance compared to stability-based Lasso-SAMBA.

Supplementary Table 3: False Discovery Rate (FDR), False Negative Rate (FNR), and $F_1$-score computed for each simulation framework, comparing the step-SAMBA, the Lasso-SAMBA, with an error control threshold of 10%, and the LassoBIC-SAMBA methods. (ND: not defined).

| **Metric** | **PK model** binomial covariates $N = 200,\ p = 500$ | **Vaccinology model** Gaussian covariates $N = 100,\ p = 200$ | **Vaccinology model** non-Gaussian covariates $N = 100,\ p = 200$ |
|---|---|---|---|
| **FDR (%) :** | | | |
| - step-SAMBA | 87.5 [78.8;91.7] | 72.7 [50.0;83.8] | 70.0 [25.0;81.2] |
| - Lasso-SAMBA | 0.0 [0.0;14.3] | 0.0 [0.0;25.0] | 0.0 [0.0;25.0] |
| - LassoBIC-SAMBA | 40.0 [0.0;64.7] | 40.0 [0.0;66.7] | 25.0 [0.0;57.1] |
| **FNR (%) :** | | | |
| - step-SAMBA | 0.0 [0.0;0.0] | 0.0 [0.0;0.0] | 0.0 [0.0;0.0] |
| - Lasso-SAMBA | 0.0 [0.0;0.0] | 0.0 [0.0;0.0] | 0.0 [0.0;0.0] |
| - LassoBIC-SAMBA | 0.0 [0.0;0.0] | 0.0 [0.0;0.0] | 0.0 [0.0;0.0] |
| **$F_1$-score (%) :** | | | |
| - step-SAMBA | 22.2 [15.4;34.9] | 42.9 [27.9;66.7] | 46.2 [31.6;85.7] |
| - Lasso-SAMBA | 100.0 [92.3;100.0] | 100.0 [85.7;100.0] | 100.0 [85.7;100.0] |
| - LassoBIC-SAMBA | 75.0 [52.2;100.0] | 75.0 [50.0;100.0] | 85.7 [60.0;100.0] |

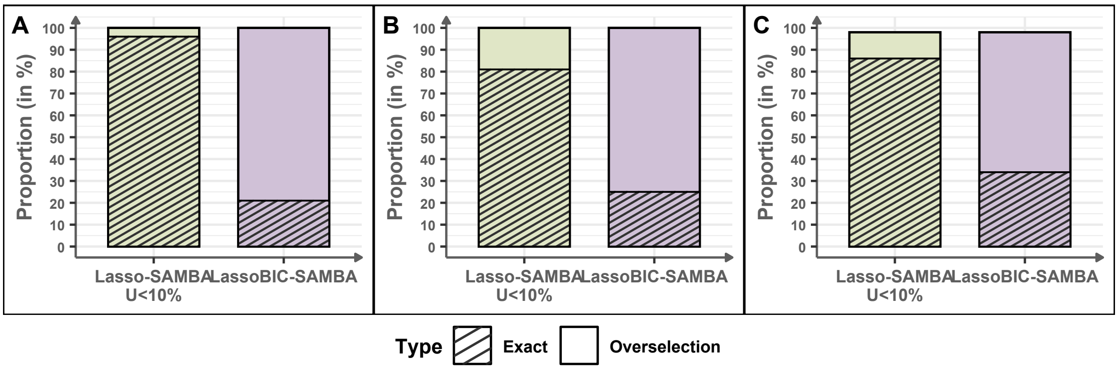

Supplementary Figure 5: Proportion of exact models (no false positives or false negatives) and models that strictly include the exact model (no false negatives but with false positives) by the Lasso-SAMBA, with an error control threshold of 10%, and the LassoBIC-SAMBA methods. (A) Pharmacokinetic model with categorical covariates ($N$ = 200, $p$ = 500); (B) Vaccinology framework with Gaussian-correlated covariates ($N$ = 100, $p$ = 200); (C) Vaccinology framework with randomly drawn correlated covariates ($N$ = 100, $p$ = 200).

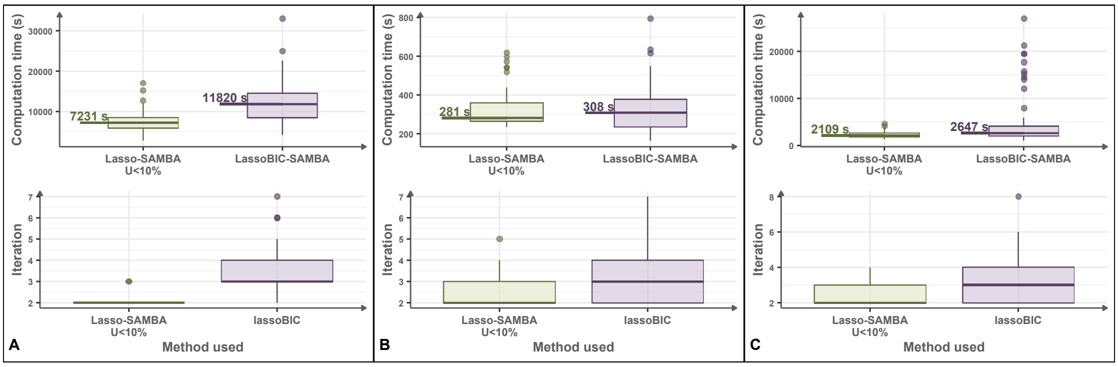

Supplementary Figure 6: Comparison of computation time distributions across simulation frameworks, for the Lasso-SAMBA, with the error control threshold set at 10%, and LassoBIC-SAMBA methods. (A) Pharmacokinetic model with categorical covariates ($N$ = 200, $p$ = 500); (B) Vaccinology framework with Gaussian-correlated covariates ($N$ = 100, $p$ = 200); (C) Vaccinology framework with randomly drawn correlated covariates ($N$ = 100, $p$ = 200).

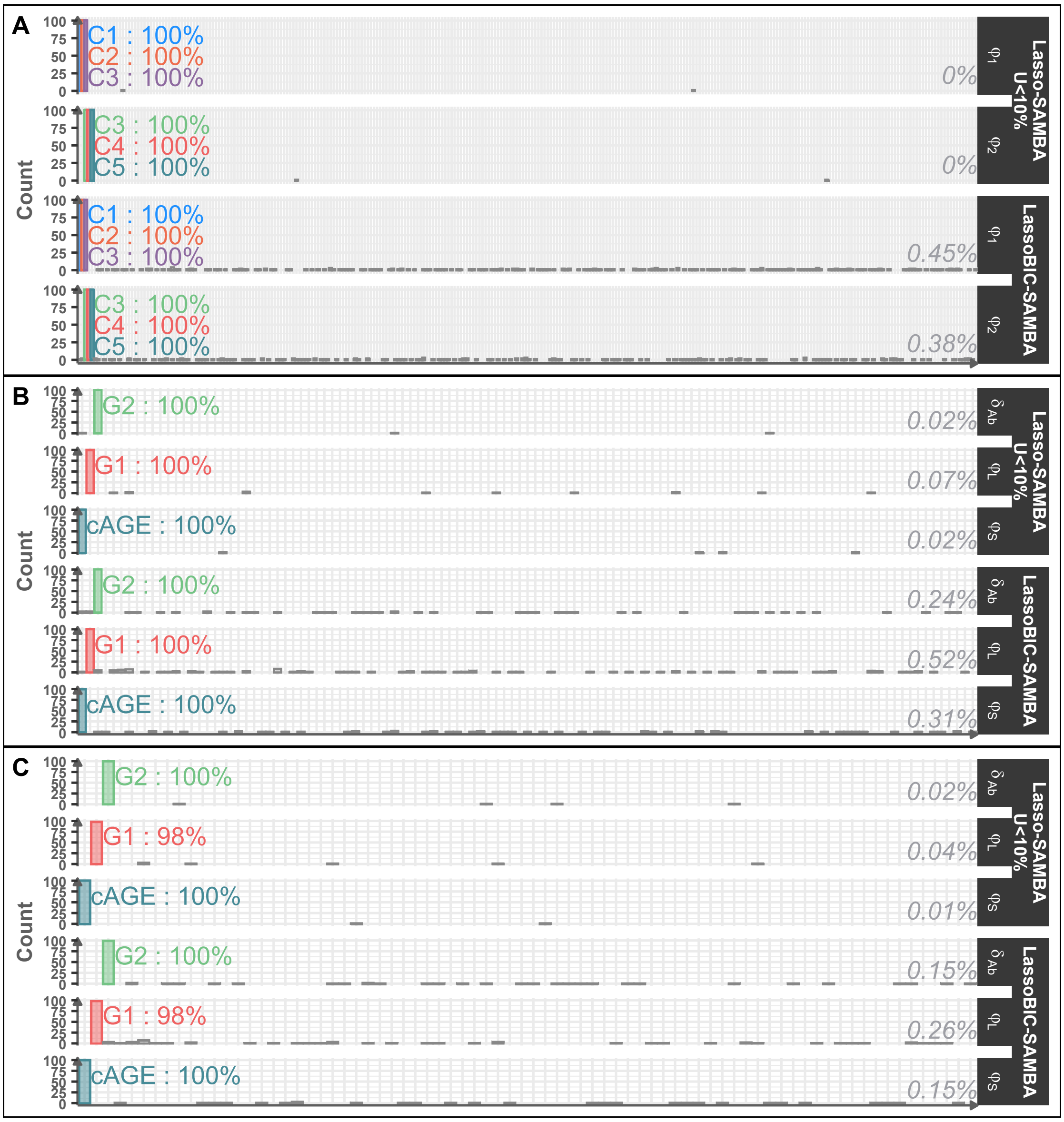

Supplementary Figure 7: Covariate selection frequency across simulation frameworks for the Lasso-SAMBA, with the error control threshold set at 10%, and the LassoBIC-SAMBA methods. The mean selection frequency of false discoveries is shown on the right side of each histogram. (A) Pharmacokinetic model with categorical covariates ($N = 200$, $p = 500$); (B) Vaccinology framework with Gaussian-correlated covariates ($N = 100$, $p = 200$); (C) Vaccinology framework with randomly drawn correlated covariates ($N = 100$, $p = 200$).

#### A.3. Multiple posterior samples of individual parameters

##### A.3.1. Method

In the following, covariate selection is again performed within the regression framework defined in Equation (S.1), linking the sampled individual parameters to the observed covariates. The specificity of this alternative lies in the way individual parameters are generated at each iteration of the algorithm.

Instead of drawing batches of individuals of size $\lfloor N/2 \rfloor$ as in the stability-enhanced procedure of Section 2.3, we repeatedly sample multiple replicates of individual parameters $(\boldsymbol{\psi}_i^{(k)})_{i\leq N}$ from their posterior distribution. Each replicate provides a full synthetic dataset of individual parameters, on which the regression model of Equation (S.1) is fitted with a Lasso penalty.

Formally, for each replicate $r \leq R$, we denote by $\hat{S}_r^\lambda$ the set of selected covariates obtained from the Lasso regression

$$(\hat{\mu}_r^\lambda, \hat{\boldsymbol{\beta}}_r^\lambda) = \underset{\mu\in\mathbb{R},\, \boldsymbol{\beta}\in\mathbb{R}^p}{\arg\min} \left\{ \sum_{i\leq N} \left( h(\psi_{il}^{(k,r)}) - \mu - \boldsymbol{X}_i \boldsymbol{\beta} \right)^2 + \lambda \|\beta\|_1 \right\}.$$

The final selection frequency for a covariate $c$ is then defined as

$$\hat{\pi}^\lambda(c) = \frac{\#\{r \leq R \mid \hat{\beta}_{c,r}^\lambda \neq 0\}}{R}.$$

As in the stability selection framework of Section 2.3, the set of selected covariates is defined as

$$\hat{\mathcal{S}}^{\lambda,\pi} = \{c \leq p \mid \hat{\pi}^\lambda(c) \geq \pi\}.$$

The calibration of $(\lambda, \pi)$ is performed following the same strategy as described in Section 2.4, i.e. exploring a grid $\Lambda \times \Pi$ and retaining the model that minimizes the Bayesian Information Criterion (BIC) while respecting the upper bound on the expected number of false discoveries [4].

This approach, later referred to as LassoREP-SAMBA, thus differs from the stability-based Lasso-SAMBA only in the construction of the replicates: by sampling multiple sets of individual parameters from the posterior distribution rather than subsampling individuals, it provides an alternative way to stabilize variable selection. Although this method yielded promising results, it comes with a much higher computational cost, since each replicate requires a complete sampling of posterior individual parameters.

##### A.3.2. Results for LassoREP-SAMBA

We compared the LassoREP-SAMBA algorithm with the stability-based Lasso-SAMBA on the vaccinology-inspired simulation frameworks with Gaussian-correlated covariates ($N = 100$, $p = 200$). Due to its high computational cost, the LassoREP-SAMBA was not applied to the pharmacokinetic or the heterogeneous vaccinology scenario.

In terms of covariate selection frequencies, as displayed in Supplementary Figure 10, both methods controlled false negatives well, with Lasso-SAMBA achieving zero false negatives and LassoREP-SAMBA showing only a single false negative across replicates. However, LassoREP-SAMBA tended to produce more false positives, particularly for covariates associated with the parameter $\varphi_S$, which were selected more frequently than with other parameters.

These trends are confirmed in the error metrics summarized in Supplementary Table 4. The median FDR of LassoREP-SAMBA reached 25.0% (95% CI: 0.0%, 57.4%), whereas Lasso-SAMBA maintained a median FDR of 0.0% (95% CI: 0.0%, 25.0%). Both methods achieved an FNR of 0.0%, and the median $F_1$-score dropped from 100.0% (95% CI: 85.7%, 100.0%) with Lasso-SAMBA to 85.7% (95% CI: 60.0%, 100.0%) with LassoREP-SAMBA.

Model selection accuracy, displayed in Supplementary Figure 8, was also reduced under LassoREP-SAMBA: the true model was recovered in only 41% of replicates, compared with 81% for Lasso-SAMBA.

The most striking difference between the two methods lies in computational efficiency, as shown in Supplementary Figure 9. The median runtime of LassoREP-SAMBA exceeded three hours (11 261 seconds), while Lasso-SAMBA required less than five minutes (281 seconds). Even in the most favorable cases, LassoREP-SAMBA took at least ten times longer than Lasso-SAMBA, with maximal runtimes exceeding 1.7 hours compared with a maximum of about 10 minutes for the baseline approach.

Overall, while LassoREP-SAMBA provides an alternative that achieves reasonable covariate recovery, its substantially higher computational burden combined with reduced accuracy makes it less attractive than the stability-based Lasso-SAMBA.

| Method | **FDR (%)** | **FNR (%)** | **F1-score (%)** |
|---|---|---|---|
| step-SAMBA | 72.7 [50.0;83.8] | 0.0 [0.0;0.0] | 42.9 [27.9;66.7] |
| Lasso-SAMBA | 0.0 [0.0;25.0] | 0.0 [0.0;0.0] | 100.0 [85.7;100.0] |
| LassoREP-SAMBA | 25.0 [0.0;57.1] | 0.0 [0.0;0.0] | 85.7 [60.0,100.0] |

Supplementary Table 4: False Discovery Rate (FDR), False Negative Rate (FNR), and $F_1$-score computed for each simulation framework, comparing the step-SAMBA, the Lasso-SAMBA and the LassoREP-SAMBA methods, with an error control threshold of 10% in the Vaccinology framework with Gaussian-correlated covariates ($N = 100$, $p = 200$).

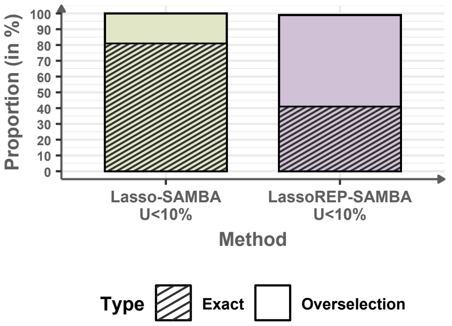

Supplementary Figure 8: Proportion of exact models (no false positives or false negatives) and models that strictly include the exact model (no false negatives but with false positives) by the Lasso-SAMBA and the LassoREP-SAMBA methods in the Vaccinology framework with Gaussian-correlated covariates ($N = 100$, $p = 200$).

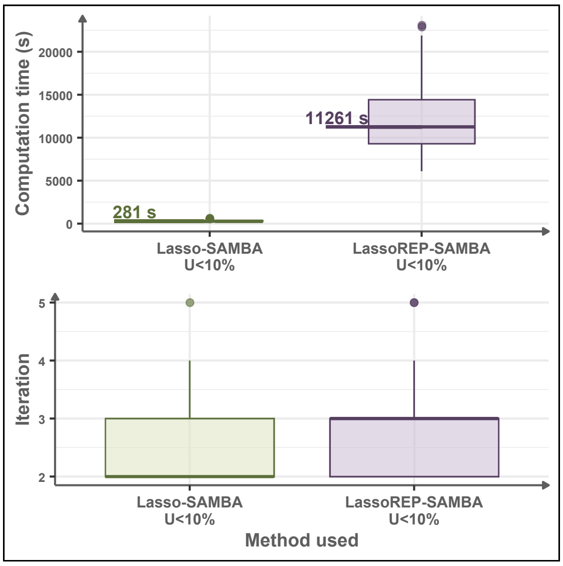

Supplementary Figure 9: Comparison of computation time distributions across simulation frameworks, for the Lasso-SAMBA and LassoREP-SAMBA methods, with the error control threshold set at 10% in the Vaccinology framework with Gaussian-correlated covariates ($N = 100$, $p = 200$).

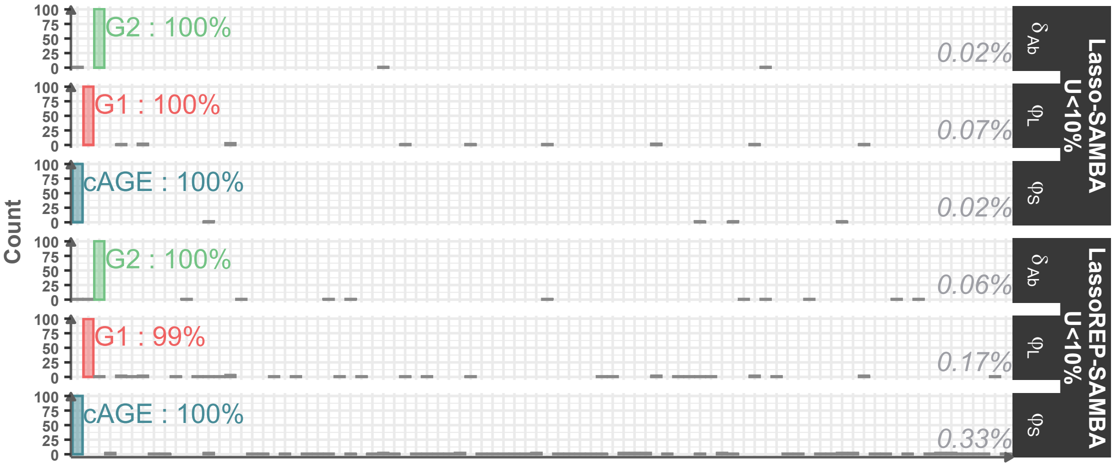

Supplementary Figure 10: Covariate selection frequency across simulation frameworks for the Lasso-SAMBA and the LassREP-SAMBA methods, with an error control threshold of 10%. The mean selection frequency of false discoveries is shown on the right side of each histogram in the Vaccinology framework with Gaussian-correlated covariates ($N = 100$, $p = 200$).

### B. Simulation generation process

For all simulated datasets, participant age, when considered, was centered following standard preprocessing practices before model fitting.

#### B.1. Pharmacokinetic model

For the growth model, we used the datasets provided by Naveau et al. [5], available at `https://github.com/Marion-Naveau/Supp_Information_SAEMVS/tree/main`. The pharmacokinetic model is defined as:

$$y = \frac{D\varphi_1}{V\varphi_1 - \varphi_2}\left(e^{-\varphi_2 t/V} - e^{-\varphi_1 t}\right),$$

where the population parameters include the dose $D$, the volume $V$, and the two rate parameters $\varphi_1$ and $\varphi_2$. Inter-individual variability is introduced on $\varphi_1$ and $\varphi_2$, such that for each individual $i \le N$:

$$\begin{cases} \varphi_{1_i} & = & \varphi_{1_{pop}} + \beta_{11}C_1 + \beta_{12}C_2 + \beta_{13}C_3 + \eta_i^1, \\ \varphi_{2_i} & = & \varphi_{2_{pop}} + \beta_{23}C_3 + \beta_{24}C_4 + \beta_{25}C_5 + \eta_i^2, \end{cases}$$

with $\boldsymbol{\eta}_i = (\eta_i^1, \eta_i^2) \overset{i.i.d.}{\sim} \mathcal{N}(0, \Omega)$.

The error model is constant, yielding the following observation equation:

$$Y_{ij} = \frac{D\varphi_{1_i}}{V\varphi_{1_i} - \varphi_{2_i}}\left(e^{-\varphi_{2_i} t/V} - e^{-\varphi_{1_i} t}\right) + \varepsilon_{ij},$$

where $\varepsilon_{ij} \overset{i.i.d.}{\sim} \mathcal{N}(0, \sigma^2)$.

Each of the $N = 200$ individuals is observed at 12 time points:

$$\{0.05, 0.15, 0.25, 0.4, 0.5, 0.8, 1, 2, 7, 12, 24, 40\}.$$

The simulation values used are summarized in Supplementary Table 5.

| Parameter | Definition | Mean value |
|---|---|---|
| **Fixed Effects** | | |
| $D$ | Dose | 100 |
| $V$ | Volume | 30 |
| $\varphi_{1_{pop}}$ | Equilibrium rate | 6 |
| $\beta_{11}$ | MF per unit increase in $C_1$ | 3 |
| $\beta_{12}$ | MF per unit increase in $C_2$ | 2 |
| $\beta_{13}$ | MF per unit increase in $C_3$ | 1 |
| $\varphi_{2_{pop}}$ | Elimination rate | 8 |
| $\beta_{23}$ | MF per unit increase in $C_3$ | 3 |
| $\beta_{24}$ | MF per unit increase in $C_4$ | 2 |
| $\beta_{25}$ | MF per unit increase in $C_5$ | 1 |
| **Random Effects** | | |
| $\Omega$ | Covariance matrix for $(\varphi_1, \varphi_2)$ | $\begin{pmatrix} 0.2 & 0.05 \\ 0.05 & 0.1 \end{pmatrix}$ |
| **Error Model** | | |
| $\sigma^2$ | Error variance | $10^{-3}$ |

Supplementary Table 5: Simulation parameters for the pharmacokinetic model based on Naveau et al. [5] (*MF: Multiplicative factor*).

##### B.2. Humoral Immune Response

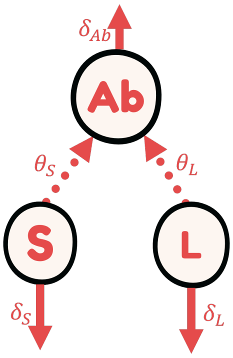

Supplementary Figure 11: Structure of the mechanistic model describing the humoral immune response.

To simulate antibody dynamics, we adopted a non-linear mixed effects model based on the work of Pasin et al. [6], previously used to characterize vaccine-induced humoral responses (Figure 11). The model integrates short-lived (S) and long-lived (L) antibody-secreting cells (ASCs), each contributing to antibody production with distinct kinetics.

- **Structural Model** :

$$\forall i \leq N, j \leq n_i, \quad \begin{cases} \frac{d}{dt}Ab_i(t_{ij}) &= \varphi_{S_i} e^{-\delta_S t_{ij}} + \varphi_{L_i} e^{-\delta_L t_{ij}} - \delta_{Ab_i} Ab_i(t_{ij}) \\ Ab_i(t_{i0} = 0) &= Ab_{0i} \end{cases}$$

$$\begin{cases} \log(\varphi_{S_i}) &= \log(\varphi_{S_{pop}}) + \log(FC\Delta AGE = +1an) \times \left(AGE_i - \overline{AGE}\right) + \eta_i^S \\ \log(\varphi_{L_i}) &= \log(\varphi_{L_{pop}}) + \beta_{G_1} \times G_{1i} + \eta_i^L \\ \log(\delta_{Ab_i}) &= \log(\delta_{Ab_{pop}}) + \beta_{G_2} \times G_{2i} + \eta_i^{Ab} \end{cases} \qquad \text{(Mod Ab Str)}$$

- **Statistical Model** :

$$\forall i \leq N, \quad \begin{cases} \eta_i^S & \overset{iid}{\sim} \mathcal{N}(0, \omega_S^2) \\ \eta_i^L & \overset{iid}{\sim} \mathcal{N}(0, \omega_L^2) \\ \eta_i^{Ab} & \overset{iid}{\sim} \mathcal{N}(0, \omega_{Ab}^2) \end{cases} \qquad \text{(Mod Ab Stat)}$$

- **Observation Model** :

$$\forall i \leq N, j \leq n_i, \quad \begin{cases} Y_{ij} & = & \log_{10}(Ab_i(t_{ij})) + \varepsilon_{ij} \\ \varepsilon_i & \overset{iid}{\sim} & \mathcal{N}(0, \Sigma = \sigma_{Ab}^2 I_{n_i}) \end{cases} \qquad \text{(Mod Ab Obs)}$$

The parameter values were based on estimates from the EBOVAC trial [7, 8, 6] and are provided in Supplementary Table 6. Antibody concentrations were simulated at time points $t_j \in \{0, 7, 21, 123, 180, 300\}$.

To create high-dimensional covariate settings, we introduced 200 covariates per individual, including 197 noise variables and 3 informative ones ($AGE$, $G_1$, $G_2$). Two covariate generation scenarios were considered: one with standard Gaussian covariates, and one with covariates drawn from randomly selected heterogeneous distributions. Details of the covariate simulation process are provided in Section B.3.

| Parameter | Definition | Value |
|---|---|---|
| **Fixed Effects** | | |
| *Influx rates* | | |
| $\varphi_S$ | ASC-S antibody production (Eu/mL/day) | 3057 at age 35 |
| $FC_{AGE}$ | Fold change per year of age | 0.934 |
| $\varphi_L$ | ASC-L antibody production (Eu/mL/day) | 16.6 when $G_1 = 0$ |
| $\beta_{G_1}$ | Effect of $G_1$ on $\varphi_L$ | 1.2 |
| *Decay rates* | | |
| $\delta_{Ab}$ | Antibody decay rate (1/day) | 0.0251 when $G_2 = 0$ |
| $\beta_{G_2}$ | Effect of $G_2$ on $\delta_{Ab}$ | 0.8 |
| $\delta_S$ | ASC-S decay rate | 0.23 |
| $\delta_L$ | ASC-L decay rate | $3.16 \cdot 10^{-4}$ |
| **Random Effects** | | |
| $\omega_S$ | SD for $\varphi_S$ | 0.92 |
| $\omega_L$ | SD for $\varphi_L$ | 0.85 |
| $\omega_{Ab}$ | SD for $\delta_{Ab}$ | 0.30 |
| **Observation Error** | | |
| $\sigma_{Ab}$ | SD of error on log antibody levels | 0.11 |

Supplementary Table 6: Parameter values for the humoral immune response model based on the EBOVAC trial [8, 6]. *(FC: fold change; SD: standard deviation)*

#### B.3. Covariate Generation

To simulate realistic dependencies among covariates, we introduced correlation structures reflecting dependencies observed in biomedical datasets. The theoretical correlation matrix used to generate the datasets in the vaccinology framework with 200 covariates is shown in Supplementary Figure 12.

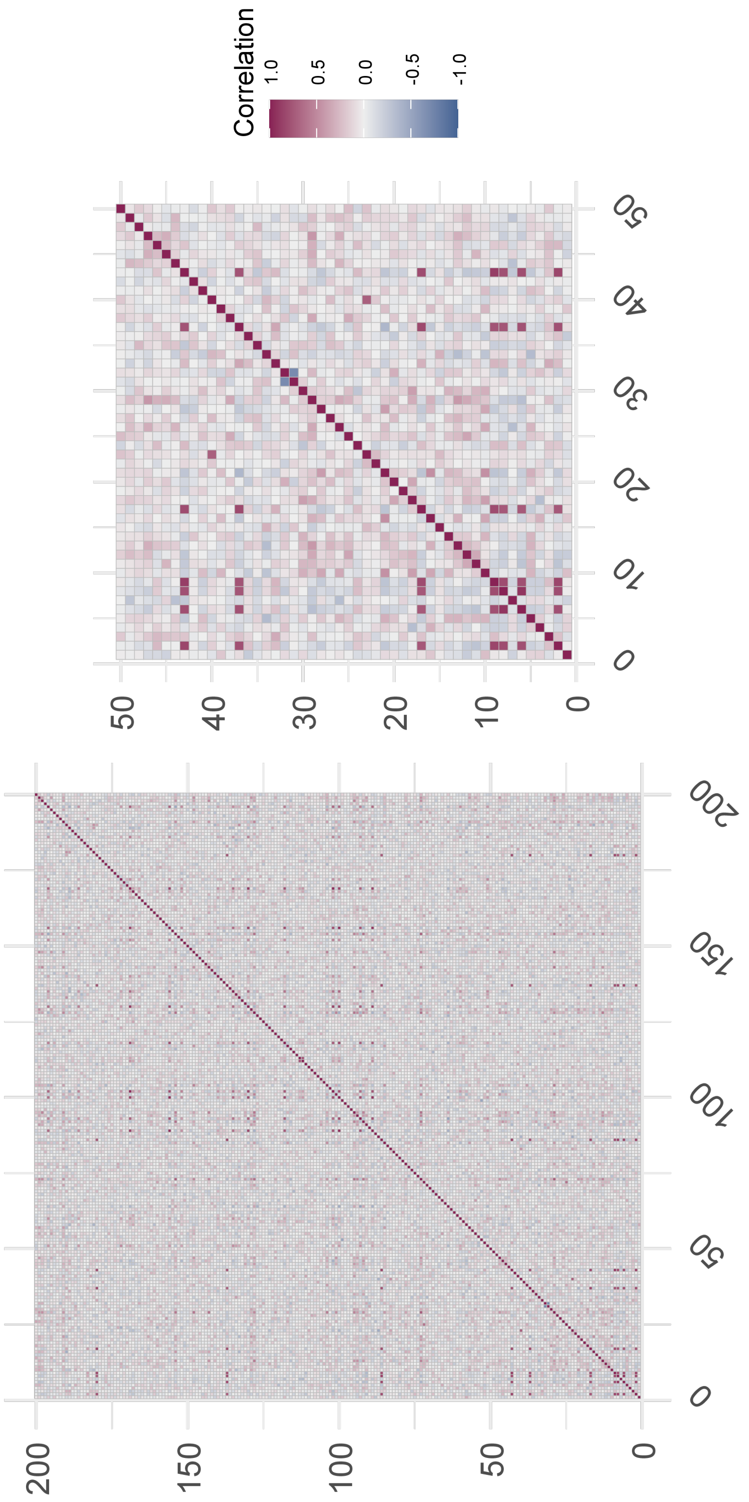

Supplementary Figure 12: Correlation matrix used to generate the 100 datasets in the vaccinology simulation framework with 200 covariates (left: full matrix; right: zoom on the first 50 covariates).

In the first scenario, we generated all covariates from a multivariate normal distribution with the specified correlation structure using the `mvtnorm` package [9] in R. In the second scenario, each covariate was drawn from a randomly selected distribution among a predefined set (Gaussian, Gamma, Poisson, Uniform). To automate this process, we implemented a function called `randomCovariate`, available on GitHub at `https://github.com/aurianegbt/LassoSAMBA`, which returns the name of a distribution and its associated parameters. The parameters are then sampled to define the distribution, and covariates are generated accordingly. Table 7 summarizes the rules used to define the parameters of each distribution.

To generate covariates with controlled correlations under the mixed-distribution scenario, we used the `simstudy` package [10]. It enables conditional simulation of variables using a flexible table-based specification.

The data definition table includes the following columns:

- `varname`: name of the variable to simulate.
- `formula`: value or R formula (defining the mean or transformation).
- `variance`: variance or another scale parameter.
- `dist`: name of the distribution (e.g., "gamma", "norm").
- `link`: link function between the formula and distribution parameters (identity, log, or logit).

Each variable is conditionally generated using the values or distributions defined above, and the resulting dataset is aligned with the specified correlation matrix using `genCorGen()` from `simstudy`. This approach ensures diverse, realistic high-dimensional covariate structures while maintaining reproducibility.

| **Suffix for the distribution** | **Element(s)** | **Generation** |
|---|---|---|
| Gamma distribution | | |
| "gamma" | "shape" | $\mathscr{P}(10)$ |
| | "scale" | $\mathscr{E}(1/2)$ |
| Normal Distribution | | |
| "norm" | "mean" | $\mathscr{N}(0,1)$ |
| | "sd" | $\mathscr{E}(0.1)$ |
| Poisson distribution | | |
| "pois" | "lambda" | $\mathscr{E}(0.1)$ |
| Uniform distribution | | |
| "unif" | | $x = \mathscr{N}(0_2, I_2)$ |
| | "min" | $\min(x)$ |
| | "max" | $\max(x)$ |

Supplementary Table 7: Process of generating various distributions using the implemented `randomCovariate` function.

### C. Additional analysis for the real-data application

#### C.1. Initial model

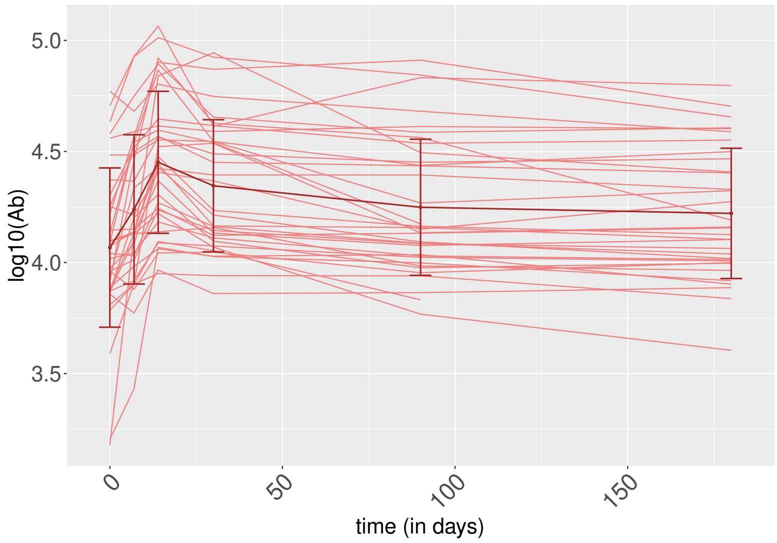

Supplementary Figure 13: Antibody titre trajectories for the 35 individuals receiving the ZOSTAVAX vaccine.

To describe the antibody kinetics observed in Supplementary Figure 13, we fitted the humoral immune response model introduced by Pasin et al. [6]. The model structure is as follows:

▸▸ **Structural Model** :

$$\forall i \leq N, j \leq n_i, \quad \begin{cases} \frac{d}{dt} Ab_i(t_{ij}) &= \varphi_{S_i} e^{-\delta_{S_i} t_{ij}} + \varphi_{L_i} e^{-\delta_L t_{ij}} - \delta_{Ab} Ab_i(t_{ij}) \\ Ab_i(t_{i0} = 0) &= Ab_{0i} \end{cases}$$

$$\begin{cases} \log(\varphi_{S_i}) &= \log(\varphi_{S_{pop}}) + \eta_i^S \\ \log(\varphi_{L_i}) &= \log(\varphi_{L_{pop}}) + \eta_i^L \\ \log(\delta_{S_i}) &= \log(\delta_{S_{pop}}) + \eta_i^\delta \end{cases} \tag{S.4}$$

▸▸ **Statistical Model** :

$$\forall i \leq N, \quad \begin{cases} \eta_i^S &\overset{iid}{\sim} \mathcal{N}(0, \omega^2_{\varphi_S}) \\ \eta_i^L &\overset{iid}{\sim} \mathcal{N}(0, \omega^2_{\varphi_L}) \\ \eta_i^\delta &\overset{iid}{\sim} \mathcal{N}(0, \omega^2_{\delta_S}) \end{cases} \tag{S.5}$$

▸▸ **Observation Model** :

$$\forall i \leq N, j \leq n_i, \quad \begin{cases} Y_{ij} & = & \log_{10}(Ab_i(t_{ij})) + \varepsilon_{ij} \\ (\varepsilon_i j)_{j \leq n_i} & \overset{iid}{\sim} & \mathcal{N}(0, \Sigma = \sigma_{Ab}^2) \end{cases} \tag{S.6}$$

To resolve identifiability issues, we fixed the decay rates $\delta_L$ (half-life for long-lived ASCs) and $\delta_{Ab}$ (half-life for antibodies), based on likelihood profile analyses shown in Supplementary Figures 14 and 15.

Figure 14 shows that for long-lived ASCs, the log-likelihood stabilizes beyond a half-life of 5 years, indicating a lack of identifiability in the data. We therefore fixed $\delta_L = 0.00019$ day$^{-1}$, corresponding to a 10-year half-life, to avoid identifiability inversion between ASC-S and ASC-L components, given their symmetrical roles in the model.

Regarding the antibody half-life, Figure 15 displays the mean values and 95% confidence intervals for fixed antibody half-life values and L-cell half-life values exceeding 5 years. The log-likelihood reaches a maximum around 11 days of half-life. We thus fixed $\delta_{Ab} = 0.063$ day$^{-1}$. The remaining model parameters were estimated via the SAEM algorithm using Monolix, and are presented in Supplementary Table 8.

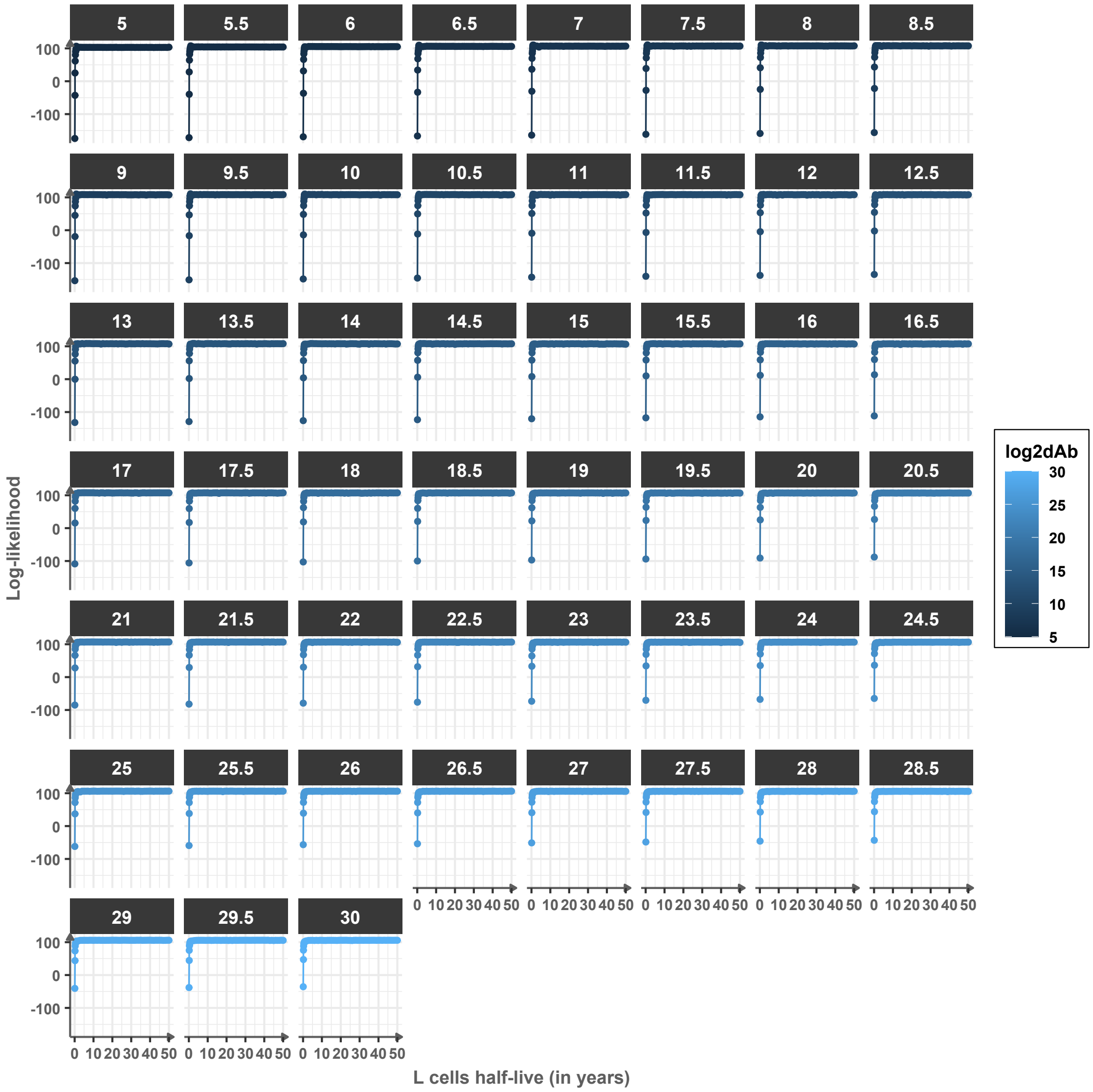

Supplementary Figure 14: Log-likelihood profile as a function of the L-cell half-life $\log(2)/\delta_L$, with antibody half-life fixed; each facet corresponds to an antibody half-life $\log(2)/\delta_{Ab}$.

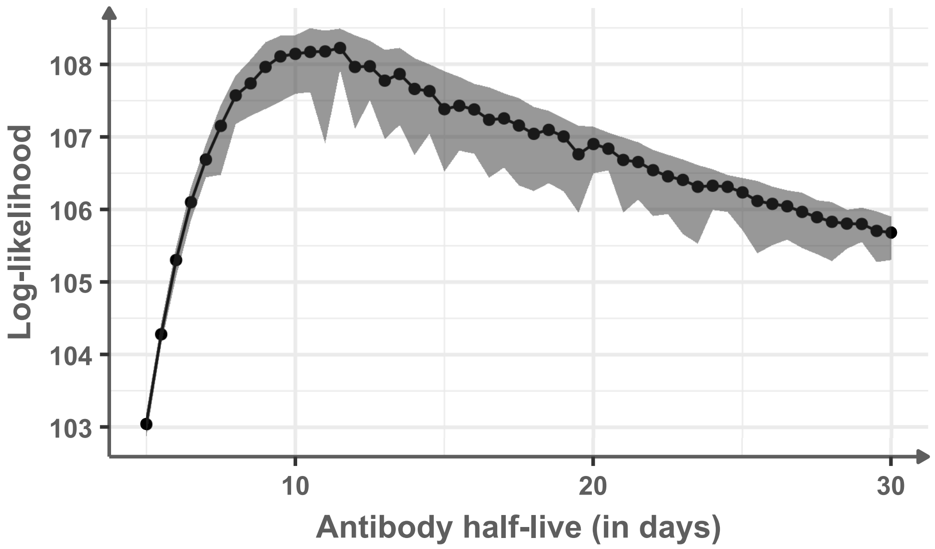

Supplementary Figure 15: Log-likelihood profile as a function of antibody half-life $\log(2)/\delta_{Ab}$. The line and ribbon indicate the mean and 95% confidence intervals over L-cell half-lives greater than 5 years.

| Parameter | Definition | Mean value (standard error) |
|---|---|---|
| **Fixed Effects** | | |
| Influx | | |
| $\varphi_{\mathbf{S_{pop}}}$ | Antibodies influx of ASC $S$ (Eu/mL/day) | 907.35 (243.66) |
| $\varphi_{\mathbf{L_{pop}}}$ | Antibodies influx of ASC $L$ (Eu/mL/day) | 1069.24 (127.66) |
| Decay rates | | |
| $\delta_{\mathbf{Ab}}$ | Decay rates of antibodies (per day) | 0.063 |
| $\delta_{\mathbf{S_{pop}}}$ | Decay rate of ASC $S$ (per day) | 0.058 (0.017) |
| $\delta_{\mathbf{L}}$ | Decay rate of ASC $L$ (per day) | $1.9 \times 10^{-4}$ |
| **Random Effects** | | |
| $\omega_{\varphi_{\mathbf{S}}}$ | Standard deviation of random effects for $\varphi_S$ | 1.16 (0.22) |
| $\omega_{\varphi_{\mathbf{L}}}$ | Standard deviation of random effects for $\varphi_L$ | 0.67 (0.084) |
| $\omega_{\delta_{\mathbf{S}}}$ | Standard deviation of random effects for $\delta_S$ | 0.50 (0.20) |
| **Error Model** | | |
| $\sigma_{\mathbf{Ab}}$ | Standard deviation of error model | 0.095 (0.0056) |

Supplementary Table 8: Estimation of the initial model parameters, without any covariates, for the ZOSTAVAX data application using the SAEM algorithm (n = 35).

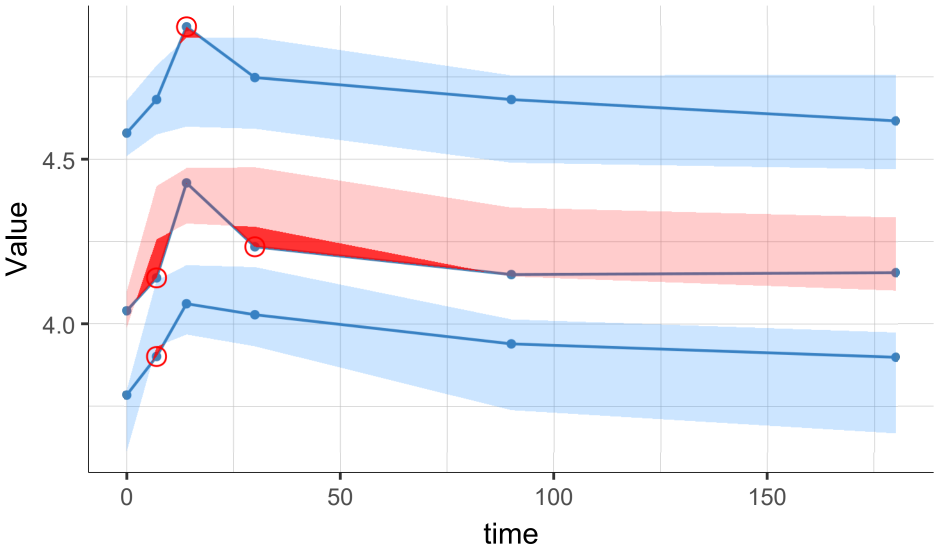

Supplementary Figure 16: Visual predictive check (VPC) of the initial model, without any covariates, for the ZOSTAVAX data application fitted using the SAEM algorithm (n = 35). The blue lines indicate the 10th, 50th, and 90th empirical percentiles; ribbons indicate the 90% prediction intervals for each percentile (10th and 90th in blue, 50th in pink). Deviations where empirical percentiles fall outside the prediction intervals are highlighted with red dots and shaded areas.

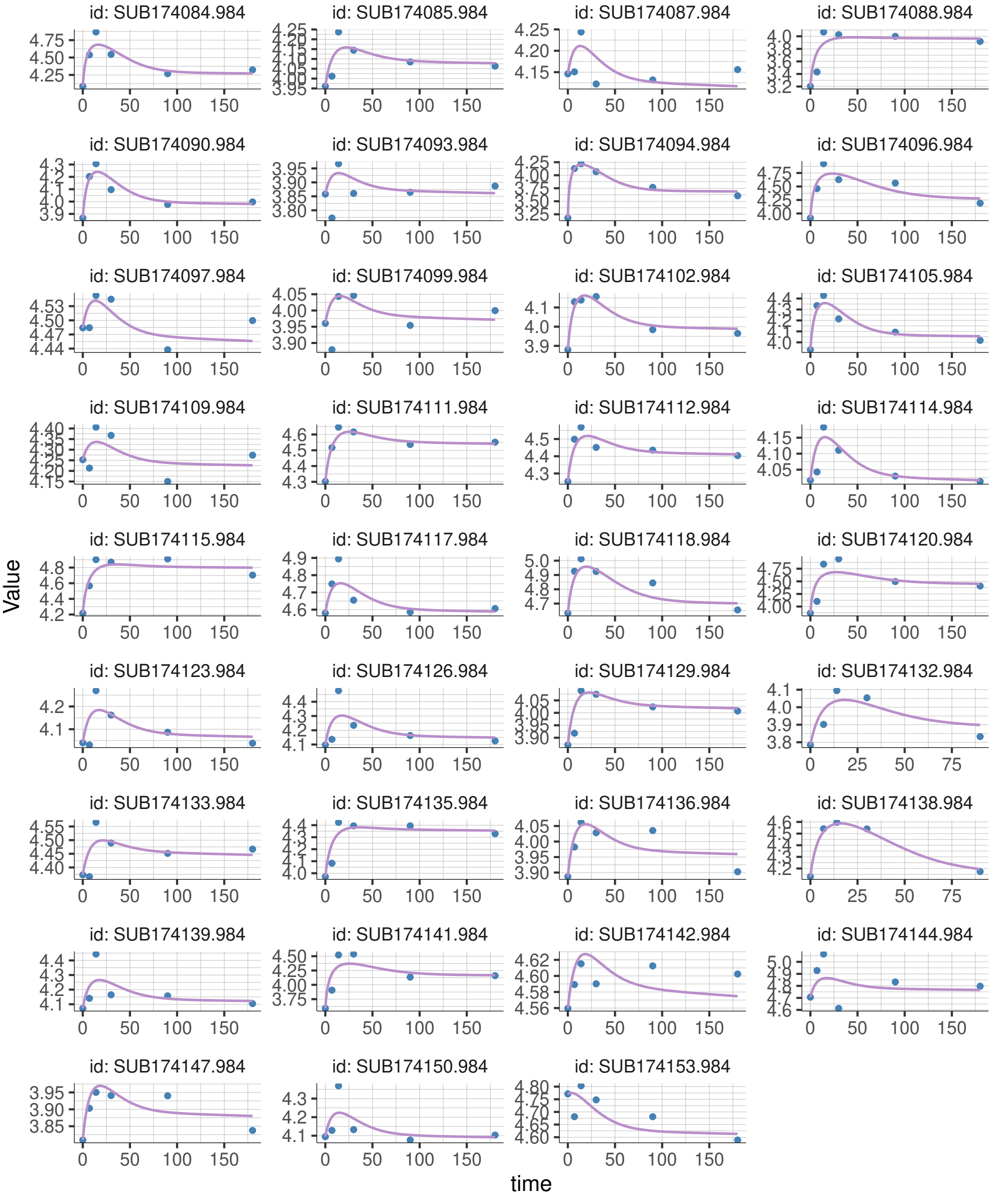

Supplementary Figure 17: Individual fits of the initial model, without any covariates, for the ZOSTAVAX data application fitted using the SAEM algorithm (n = 35). For each subject (facet), blue dots indicate observations and the purple line shows model predictions.

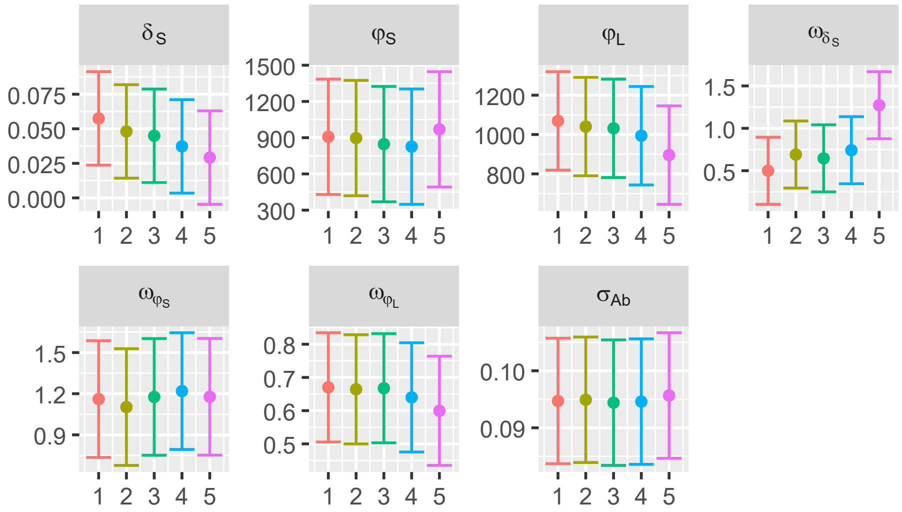

Supplementary Figure 18: Convergence diagnostics for the initial model, without any covariates, for the ZOSTAVAX data application: population parameter estimates across 5 independent Monolix runs of the SAEM algorithm with 95% confidence intervals.

#### C.2. Final Model

The final model was obtained using the Lasso-SAMBA procedure, which selected two genes: **LEP**, associated with $\varphi_S$, and **KIFC1**, associated with $\varphi_L$:

$$\log(\varphi_{S_i}) = \log(\varphi_{S_{pop}}) + \beta_{LEP} \times X_{LEP_i} + \eta_i^S,$$

and

$$\log(\varphi_{L_i}) = \log(\varphi_{L_{pop}}) + \beta_{KIFC1} \times X_{KIFC1_i} + \eta_i^L.$$

Parameters were re-estimated via SAEM in Monolix and are summarized in Supplementary Table 9. Gene expression data were centered before estimation. Figure 19 shows the antibody data stratified by gene expression levels.

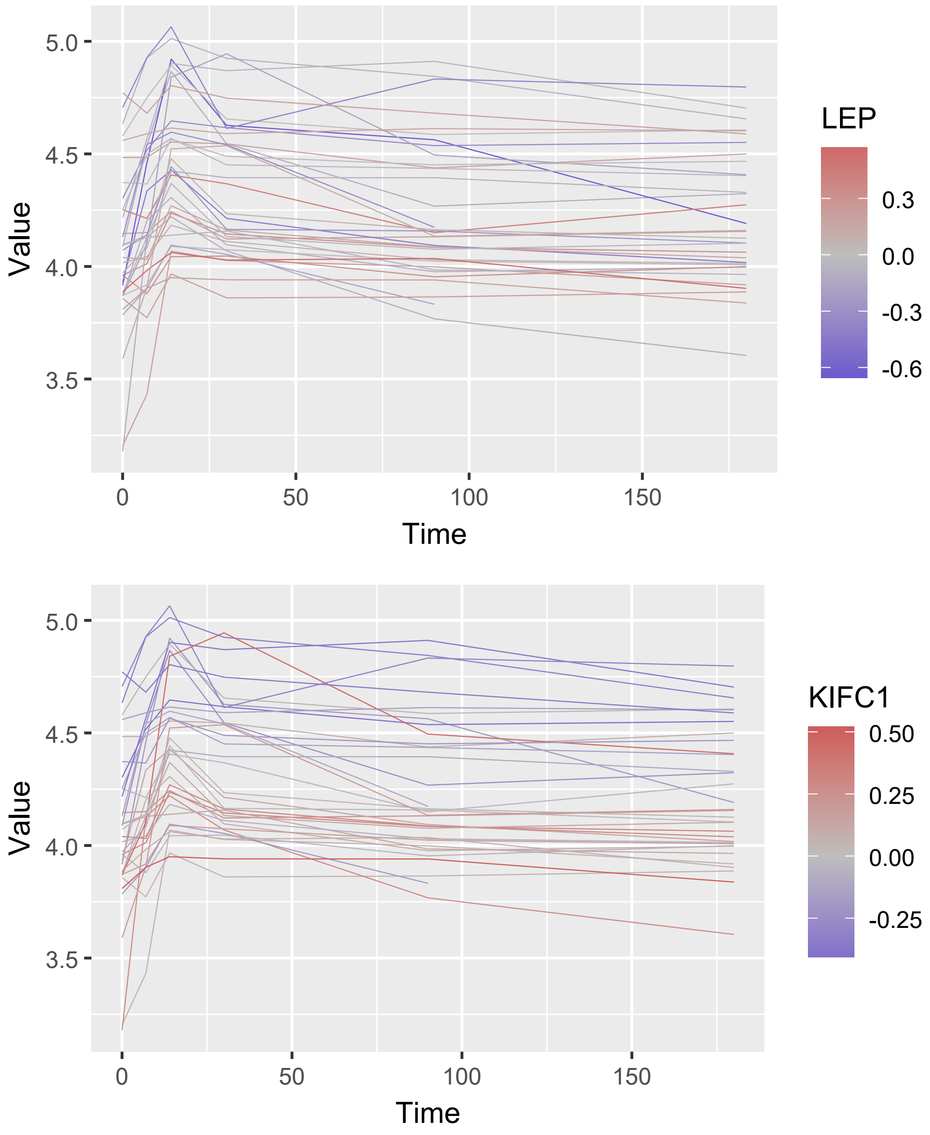

Supplementary Figure 19: Antibody trajectories of the 35 individuals vaccined with ZOSTAVAX stratified by gene expression levels measured at day 0 of LEP (top) and KIFC1 (bottom).

| Parameter | Definition | Mean value (standard error) |
|---|---|---|
| **Fixed Effects** | | |
| Influx | | |
| $\varphi_{\mathbf{S_{pop}}}$ | Antibodies influx of ASC $S$ (Eu/mL/day) | 945.43 (193.78) |
| $\beta_{\mathbf{LEP}}$ | | -3.40 (0.71) |
| $\varphi_{\mathbf{L_{pop}}}$ | Antibodies influx of ASC $L$ (Eu/mL/day) | 1007.21 (105.63) |

| | | |
|---|---|---|
| $\beta_{\mathbf{KIFC1}}$ | | -2.05 (0.43) |
| Decay rates | | |
| $\delta_{\mathbf{Ab}}$ | Decay rates of antibodies (per day) | 0.063 |
| $\delta_{\mathbf{Spop}}$ | Decay rate of ASC $S$ (per day) | 0.047 (0.018) |
| $\delta_{\mathbf{L}}$ | Decay rate of ASC $L$ (per day) | $1.9 \times 10^{-4}$ |
| **Random Effects** | | |
| $\omega_{\varphi_{\mathbf{S}}}$ | Standard deviation of random effects for $\varphi_S$ | 0.83 (0.15) |
| $\omega_{\varphi_{\mathbf{L}}}$ | Standard deviation of random effects for $\varphi_L$ | 0.66 (0.089) |
| $\omega_{\delta_{\mathbf{S}}}$ | Standard deviation of random effects for $\delta_S$ | 0.72 (0.27) |
| **Error Model** | | |
| $\sigma_{\mathbf{Ab}}$ | Standard deviation of error model | 0.094 (0.0056) |

Supplementary Table 9: Estimation of the final model parameters for the ZOSTAVAX data application using the Lasso-SAMBA method (n = 35).

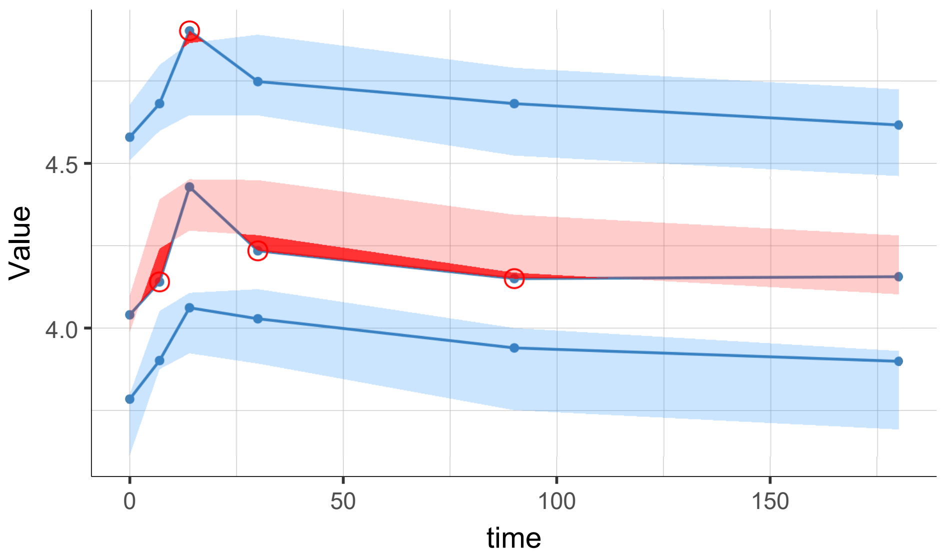

Supplementary Figure 20: Visual predictive check (VPC) of the final model built with the lasso-SAMBA method for the ZOSTAVAX data application fitted using the SAEM algorithm (n = 35). The blue lines indicate the 10th, 50th, and 90th empirical percentiles; ribbons indicate the 90% prediction intervals for each percentile (10th and 90th in blue, 50th in pink). Deviations where empirical percentiles fall outside the prediction intervals are highlighted with red dots and shaded areas.

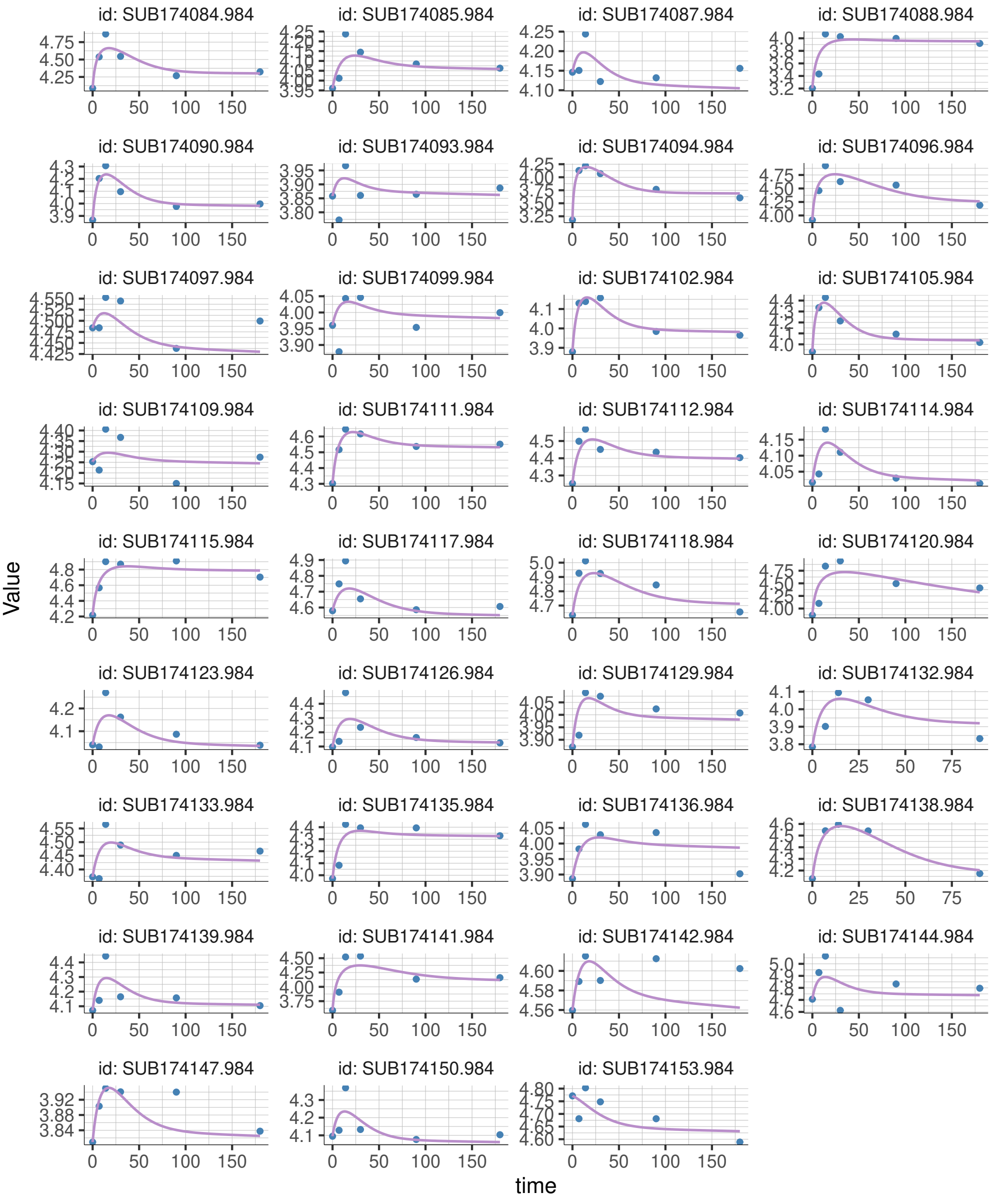

Supplementary Figure 21: Individual fits of the final model built with the lasso-SAMBA method for the ZOSTAVAX data application fitted using the SAEM algorithm (n = 35). For each subject (facet), blue dots indicate observations and the purple line shows model predictions.

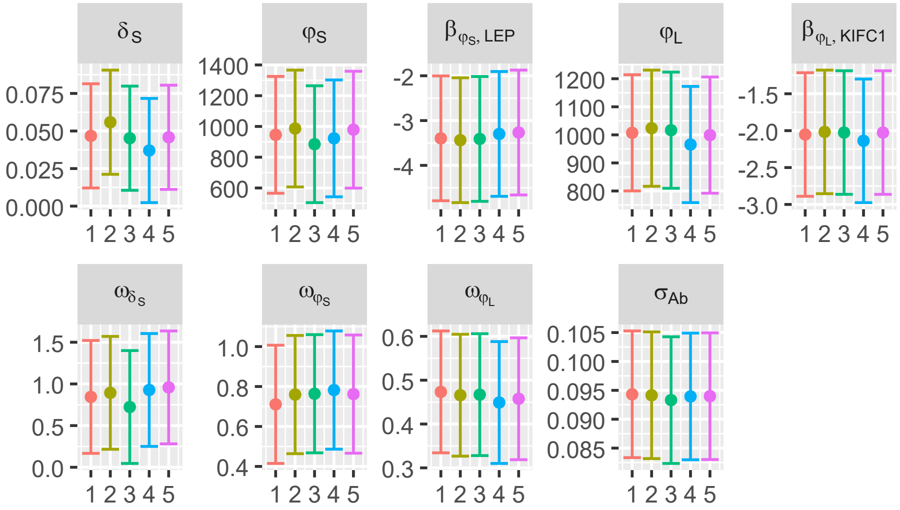

Supplementary Figure 22: Convergence diagnostics for the final model built with the lasso-SAMBA method for the ZOSTAVAX data application: population parameter estimates across 5 independent Monolix runs of the SAEM algorithm with 95% confidence intervals.

### D. Additional graphs for results

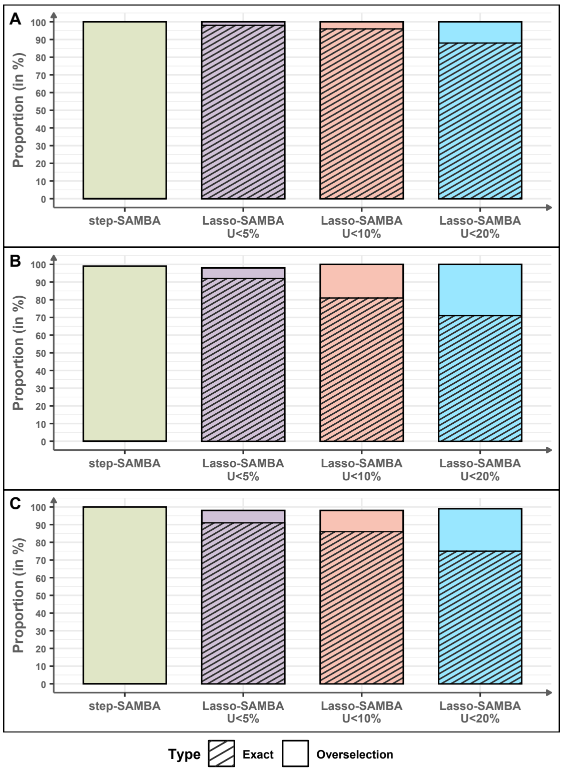

Supplementary Figure 23: Proportion of exact models (no false positives or false negatives) and models that strictly include the exact model (no false negatives but with false positives) for step-SAMBA and Lasso-SAMBA under different control thresholds. (A) Pharmacokinetic model with categorical covariates ($N$ = 200, $p$ = 500); (B) Vaccinology framework with Gaussian-correlated covariates ($N$ = 100, $p$ = 200); (C) Vaccinology framework with randomly drawn correlated covariates ($N$ = 100, $p$ = 200).

| Metric | **PK model** binomial covariates $N = 200,\ p = 500$ | **Vaccinology model** Gaussian covariates $N = 100,\ p = 200$ | **Vaccinology model** non-Gaussian covariates $N = 100,\ p = 200$ |
|---|---|---|---|
| **FDR (%) :** | | | |
| - step-SAMBA | 87.5 [78.8;91.7] | 72.7 [50.0;83.8] | 70.0 [25.0;81.2] |
| - Lasso-SAMBA $U_{\lambda,\pi} \leq 0.05$ | 0.0 [0.0;0.0] | 0.0 [0.0;25.0] | 0.0 [0.0;25.0] |
| - Lasso-SAMBA $U_{\lambda,\pi} \leq 0.1$ | 0.0 [0.0;14.3] | 0.0 [0.0;25.0] | 0.0 [0.0;25.0] |
| - Lasso-SAMBA $U_{\lambda,\pi} \leq 0.2$ | 0.0 [0.0;14.3] | 0.0 [0.0;40.0] | 0.0 [0.0;25.0] |
| - SAEMVS | 0.0 [0.0;100.0] | NA | NA |
| **FNR (%) :** | | | |
| - step-SAMBA | 0.0 [0.0;0.0] | 0.0 [0.0;0.0] | 0.0 [0.0;0.0] |
| - Lasso-SAMBA $U_{\lambda,\pi} \leq 0.05$ | 0.0 [0.0;0.0] | 0.0 [0.0;0.0] | 0.0 [0.0;0.0] |
| - Lasso-SAMBA $U_{\lambda,\pi} \leq 0.1$ | 0.0 [0.0;0.0] | 0.0 [0.0;0.0] | 0.0 [0.0;0.0] |
| - Lasso-SAMBA $U_{\lambda,\pi} \leq 0.2$ | 0.0 [0.0;0.0] | 0.0 [0.0;0.0] | 0.0 [0.0;0.0] |
| - SAEMVS | 0.0 [0.0;100.0] | NA | NA |
| **F1-score (%) :** | | | |
| - step-SAMBA | 22.2 [15.4;34.9] | 42.9 [27.9;66.7] | 46.2 [31.6;85.7] |
| - Lasso-SAMBA $U_{\lambda,\pi} \leq 0.05$ | 100.0 [100.0;100.0] | 100.0 [85.7;100.0] | 100.0 [85.7;100.0] |
| - Lasso-SAMBA $U_{\lambda,\pi} \leq 0.1$ | 100.0 [92.3;100.0] | 100.0 [85.7;100.0] | 100.0 [85.7;100.0] |
| - Lasso-SAMBA $U_{\lambda,\pi} \leq 0.2$ | 100.0 [92.3;100.0] | 100.0 [75.0;100.0] | 100.0 [85.7;100.0] |
| - SAEMVS | 100.0 [0.0;100.0] | NA | NA |

Supplementary Table 10: False Discovery Rate (FDR), False Negative Rate (FNR), and $F_1$-score computed for each simulation framework, comparing the original step-SAMBA method and the Lasso-SAMBA method under different error control thresholds. (NA: not applied).

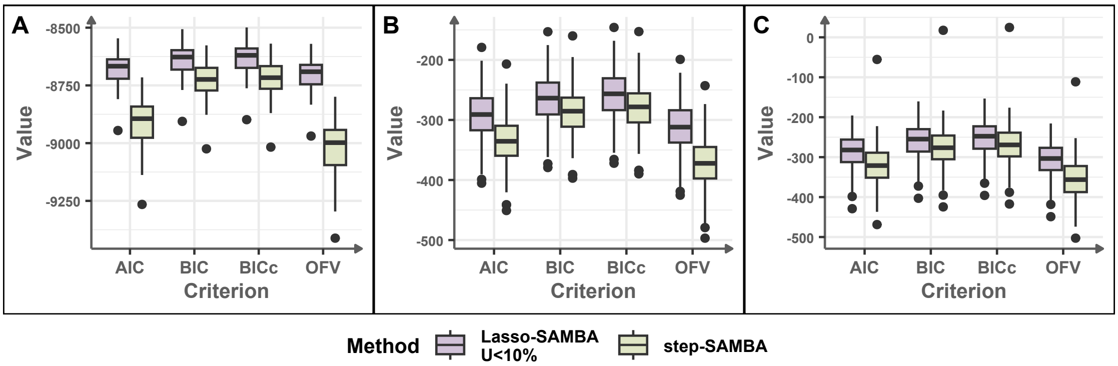

Supplementary Figure 24: Distribution of final model selection scores (AIC, BIC, BICc, OFV) across replicates in each framework, comparing step-SAMBA and Lasso-SAMBA with the error control threshold set at 10%. (A) Pharmacokinetic model with categorical covariates ($N = 200$, $p = 500$); (B) Vaccinology framework with Gaussian-correlated covariates ($N = 100$, $p = 1000$); (C) Vaccinology framework with randomly drawn correlated covariates ($N = 100$, $p = 200$)

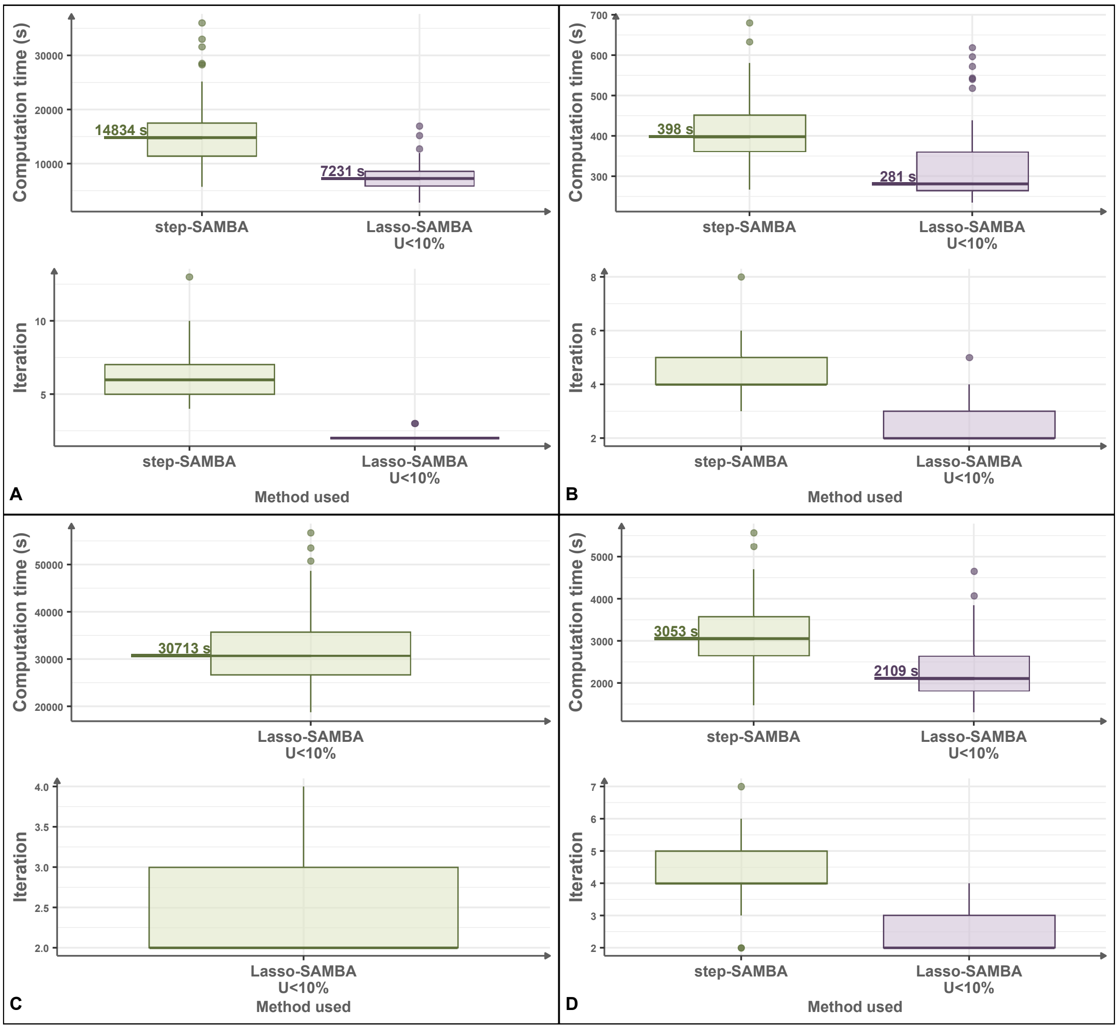

Supplementary Figure 25: Comparison of computation time distributions across simulation frameworks, for step-SAMBA and Lasso-SAMBA with the error control threshold set at 10%. (A) Pharmacokinetic model with categorical covariates ($N = 200$, $p = 500$); (B) Vaccinology framework with Gaussian-correlated covariates ($N = 100$, $p = 200$); (C) Vaccinology framework with Gaussian-correlated covariates ($N = 100$, $p = 1000$); (D) Vaccinology framework with randomly drawn correlated covariates ($N = 100$, $p = 200$)

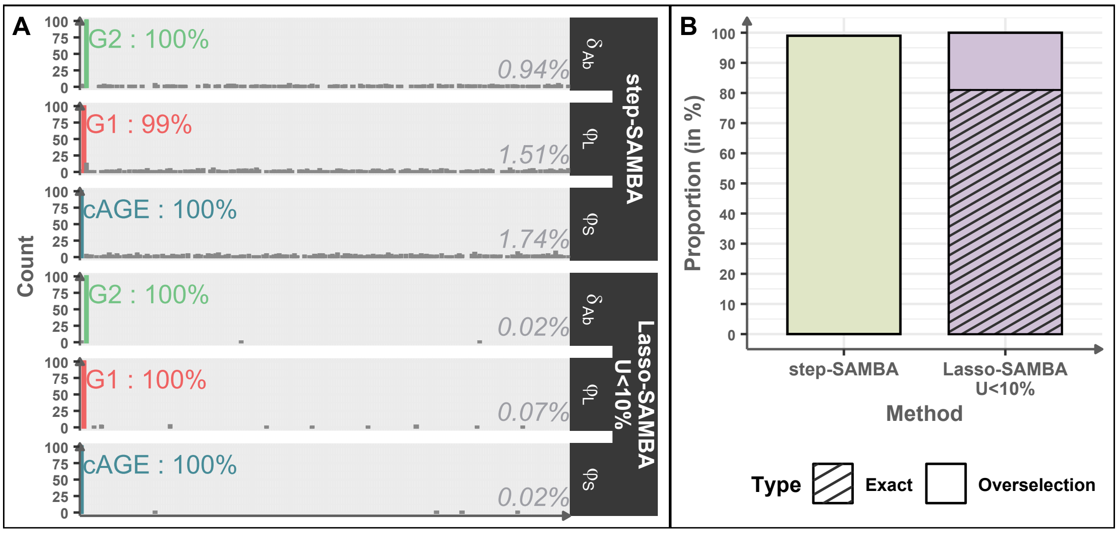

Supplementary Figure 26: Simulation results for the vaccinology framework with Gaussian-correlated covariates ($N$ = 100, $p$ = 200) for the step-SAMBA and the Lasso-SAMBA with an error control threshold of 10%. (A) Covariate selection frequency across simulation frameworks. The mean selection frequency of false discoveries is shown on the right side of each histogram. (B) Proportion of exact models (no false positives or false negatives) and models that strictly include the exact model (no false negatives but with false positives).

### References

[1] R Core Team. *R: A Language and Environment for Statistical Computing.* R Foundation for Statistical Computing, Vienna, Austria, 2024.

[8] Marie Alexandre, Mélanie Prague, Chelsea McLean, Viki Bockstal, Macaya Douoguih, Rodolphe Thiébaut, Thierry Effelterre, Laura Solforosi, and Anna Dari. Prediction of long-term humoral response induced by the two-dose heterologous Ad26.ZEBOV, MVA-BN-Filo vaccine against Ebola. *NPJ Vaccines*, 8, 11 2023.

[9] Alan Genz and Frank Bretz. *Computation of Multivariate Normal and t Probabilities.* Lecture Notes in Statistics. Springer-Verlag, Heidelberg, 2009.

[10] Keith Goldfeld and Jacob Wujciak-Jens. simstudy: Illuminating research methods through data generation. *J Open Source Softw*, 5(54):2763, 2020.
